# Mortality in Rural Coastal Kenya measured using the Kilifi Health and Demographic Surveillance System: A 16-year descriptive analysis

**DOI:** 10.1101/2021.09.16.21263698

**Authors:** Mark Otiende, Evasius Bauni, Amek Nyaguara, David Amadi, Christopher Nyundo, Emmanuel Tsory, David Walumbe, Michael Kinuthia, Norbert Kihuha, Michael Kahindi, Gideon Nyutu, Jennifer Moisi, Amare Deribew, Ambrose Agweyu, Kevin Marsh, Benjamin Tsofa, Philip Bejon, Christian Bottomley, Thomas N. Williams, J. Anthony G. Scott

## Abstract

**Background:** The Kilifi Health and Demographic Surveillance System (KHDSS) was established in 2000 to define the incidence and prevalence of local diseases and evaluate the impact of community-based interventions. KHDSS morbidity data have been reported comprehensively but mortality has not been described. This analysis describes mortality in the KHDSS over 16 years.

**Methods:** We calculated mortality rates from 2003-2018 in four intervals of equal duration and assessed differences in mortality across these intervals by age and sex. We calculated the period survival function and median survival using the Kaplan-Meier method and mean life expectancies using abridged life tables. We estimated trend and seasonality by decomposing a time series of monthly mortality rates. We used choropleth maps and random effects Poisson regression to investigate geographical heterogeneity.

**Results:** Mortality declined by 36% overall between 2003-2018 and by 59% in children aged <5 years. Most of the decline occurred between 2003 and 2006. Among adults, the greatest decline (49%) was observed in those aged 15-54 years. Life expectancy at birth increased by 12 years. Females outlived males by 6 years. Seasonality was only evident in the 1-4 year age group in the first four years. Geographical variation in mortality was +/-10% of the median value and did not change over time.

**Conclusions:** Between 2003-2018 mortality among children and young adults has improved substantially. The steep decline in 2003-2006 followed by a much slower reduction thereafter suggests improvements in health and wellbeing have plateaued in the last 12 years. However, there is substantial inequality in mortality experience by geographical location.

## BACKGROUND

A majority of Low- and Middle-Income Countries (LMICs), especially in sub-Saharan Africa (sSA), lack comprehensive Civil Registration and Vital Statistics Systems (CRVS) necessary for monitoring mortality (1, 2). Tracking the progress in child and adult survival, therefore, relies on alternative data sources such as Demographic and Health Surveys (DHS), National Population Censuses, and Health and Demographic Surveillance Systems (HDSSs). HDSSs are designed to monitor a small sub-population of a nation in a defined geographical area and are a commonly used resource for health and demographic research in LMICs.

The Kilifi Health and Demographic Surveillance System (KHDSS) was established in 2000 by the KEMRI-Wellcome Trust Research Programme (KWTRP) to monitor mortality and morbidity caused by common diseases and to provide a sampling frame for epidemiological studies (3). The surveillance area was selected to capture at least 80% of patients admitted to the Kilifi County Hospital and over the subsequent two decades, the platform has been used to describe morbidity in children and adults. This includes incidence of malaria (4), pneumonia (5), lower respiratory tract infections (6), rotavirus (7), malnutrition (8, 9), sickle cell disease (10), epilepsy (11) as well as the burden of mental health problems (12), pregnancy-related disorders and chronic diseases that contribute substantially to overall mortality (13, 14). The KHDSS has also been used to evaluate the impact of new community-based interventions such as vaccines and bed net use (5, 15-19).

Morbidity data have been reported comprehensively in Kilifi but mortality in this population has not been systematically described. In this paper, we describe mortality in children and adults over a 16-year period and analyse deaths by age, sex, season, geographical location, and temporal trend.

## METHODS

### Data source and setting

The KHDSS surveillance area, which is located in Kilifi County, within the former Coast Province, is divided into 15 government administrative regions called locations, comprising a total of 891 km^2^ (Figure S1). An initial census and mapping of the surveillance area was conducted in 2000 and was found to contain 189,148 residents in 20,978 households. Subsequently, the population has been continuously monitored for births, pregnancies, deaths, and migration events in re-enumeration rounds occurring approximately every 4 months, and the mapping was updated in 2017. At the end of 2018, there were 299,471 residents living in 41,536 households. From 2008, deaths registered in the KHDSS have also been investigated for the cause of death by Verbal Autopsy (VA) which has been reported separately (20). Kilifi County Hospital (KCH) is located at the geographical centre of the KHDSS area and, during the study period, it was the only government facility offering inpatient care for the KHDSS population. A small number of private hospitals and lower-level facilities have a few in-patient beds.

The concept of the KHDSS is based on the INDEPTH (International Network for the Demographic Evaluation of Populations and Their Health) data model (21, 22). Demographic and health data are collected at 4 points of contact: at re-enumeration when community interviewers make household visits to update the population register; at the inpatient wards of KCH where medical staff record patient history, clinical examination and outcome (death or discharge); at the maternity ward of KCH where staff record births and perinatal deaths; and in 34 vaccination clinics distributed across the surveillance area which collected data on childhood vaccination between 2008-2018. The eligibility for inclusion, the variables routinely measured, the structure of the KHDSS databases and the population structure have all been described previously (3)

Initially, data collection during household visits was paper-based but switched to electronic data collection using tablets in 2016. The tablets are loaded daily with the most recent copy of the residents’ database and, after data collection, they are returned to the research unit where a two-way synchronization with the master database is performed. All other data collection points are linked in real-time to the master database that has been specified using MySQL.

At re-enumeration rounds, information on all household members is sought from a single informant, usually a member of the household. If all household members are unavailable during the visit, information is obtained from neighbouring households. All field staff are debriefed on the quality of data collected after each enumeration cycle and re-trained where needed. The data collection applications are programmed with skip patterns and consistency checks to ensure mandatory information is collected. Additionally, within the database, there are built-in checks for missing or duplicated data.

To explore the accuracy of age data at the first census and among all new in-migrants, we calculated Whipple’s Index (23). Whipple’s index measures the tendency for individuals to inaccurately report their age in rounded numbers, usually ending in 0 and 5, resulting in age heaping.

### Statistical analysis

The analysis period, from 1 Jan 2003 to 31 Dec 2018, was stratified into 4 non-overlapping periods each lasting 4 years. We excluded 2000-2002 because of changes in the re-enumeration protocols designed to increase the ascertainment of deaths in neonates during these years. We used survival analysis and routine demographic life table methods to calculate mortality rates and life expectancy and examined seasonality, short and long-term trends over the 16 years.

#### Age-sex mortality profile

The mortality rate is calculated as the number of deaths divided by person years of observation (PYO). Entry to risk begins at the latest of birth, in-migration or study start date. Exit from risk is at the earliest of study end-date, out-migration or death. If an out-migration is followed by an in-migration, the period between the out-migration and in-migration is excluded from the risk period to avoid survivor bias. The total PYO was computed for different age groups, sex, and locations.

For children aged less than five years, we have also calculated conventional mortality ratios where the number of deaths within a specific age group in a given time period are divided by the number of live births occurring during the same time period. Mortality ratios are commonly used in settings where risk time cannot be quantified. They can be confounded by varying birth rates as the deaths in the numerator are not always drawn from the denominator population.

#### Survival and life-expectancy

We used two methods to estimate life expectancy; the period life table method which calculates the *mean* life expectancy at birth and the Kaplan-Meier (KM) survival method which calculates the *median* age at death. The main difference between the methods is in the age intervals used; the life table method computes survival probabilities within pre-define age intervals e.g. 5-year intervals whereas the KM method computes survival probabilities whenever there is a death in the cohort making the KM intervals smaller and of variable length (24).

For purposes of comparison with other analyses, we also generated abridged life tables using data structured according to analytic methods developed by the Multi-centre Analysis of the Dynamics of Internal Migration and Health (MADIMAH) which was a working group within INDEPTH. In the MADIMAH method (25, 26), the definition of risk time considers the time between out-migration and a subsequent in-migration. If the difference is less than 180 days, this time is included in the risk period which increases the person-years of observation resulting in lower estimates of mortality rates.

#### Seasonality and trend

We first assessed seasonality and long-term temporal trends for each age group by graphically reviewing a time series of monthly mortality rates. We then estimated trend and seasonality based on an STL (**S**easonal and **T**rend using **L**OESS) decomposition and identified months with the highest and lowest mortality rates from the seasonal component (27).

#### Geographical heterogeneity in survival and mortality over time

We produced choropleth maps for overall and age-specific mortality rates in the four 4-year periods to investigate the geographical variation of mortality in the administrative locations over time. For overall mortality, we accounted for temporal differences in the population age-sex structure by direct standardization against the 2011 KHDSS age-sex structure. All the maps were created at the administrative location level.

We used the quantile method to create 5 mortality rate classes for map reading. This method places equal numbers of data units (death rates) in each class resulting in classes centred on the median death rate. For each age group, the quintiles are derived from the entire mortality rate range between 2003-2018 and the resulting classification is applied across each of the 4-year-period maps for that age group. The quantile method, though simple, has been shown to be the optimal classification method for displaying geographically varying data in series in a map reading experiment [24].

To assess geographical variation in mortality, for each period, we fitted a multi-level Poisson regression model adjusting for sex and age in which location was included as a random effect and used the variance of the random effect to quantify heterogeneity. We tested for between-location variation within each period using the likelihood ratio test and also tested for temporal variation in mortality rates between the 2003-06 period and each of the subsequent periods using a z-test. We also calculated the median age at death for each of the 15 administrative locations in the 4-year periods and assessed variation in life expectancy by location and time.

All analyses were conducted using STATA/IC version 15.1 (StataCorp College Station, Texas, USA) and R version 4.1.0 (28).

## RESULTS

### Age-sex profile

The cohort consisted of 699,841 individuals of whom 125,587 (18%) were followed from birth. In total, we observed 22,207 deaths in 3,897,529 person-years. More than 95% of the information on residence and vital status was collected from respondents living in the same household. Females contributed 53% of the total PYO and 48% of deaths (Table 1). There was no indication of age heaping or misspecification of sex (Table S1).

**Table 1:**
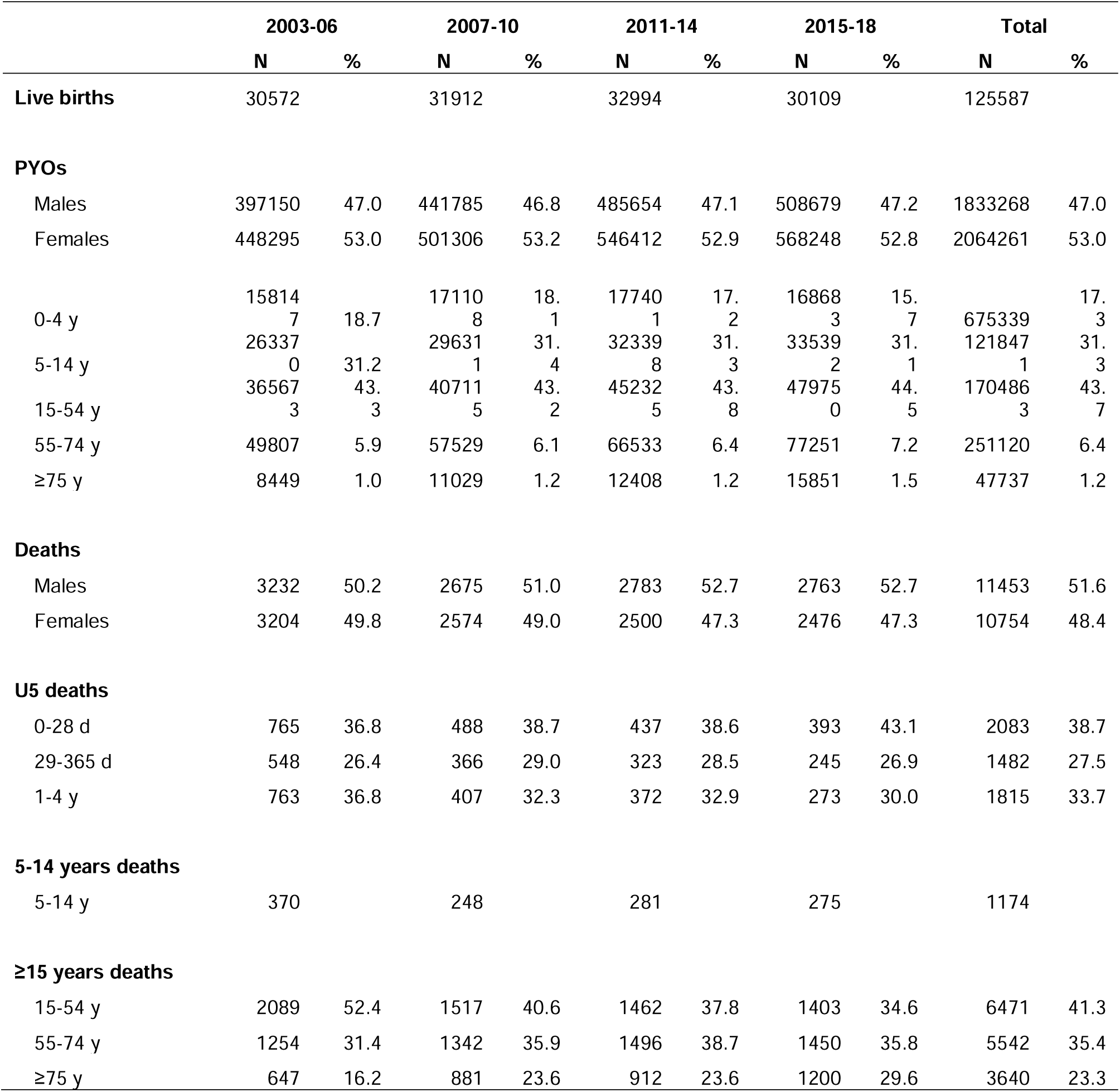
Distribution of births, deaths, and person-years by sub-period

Mortality was highest in the first time period (2003-2006) compared to the other time periods for all ages except adults aged >54 years (Figure S2a). The mortality rate in children aged <5 years declined by 44% between the periods 2003-2006 and 2007-2010, from 13.1 to 7.4 deaths per 1000 PYO, and continued to decline more slowly reaching 5.4 in 2015-2018; we observed a similar pattern in all the finer age-strata of children aged <5 years (Table 2). Mortality ratios, per 1000 live births, for children aged <5 years, are presented in Table S2. Over the whole 16-year period mortality rates were lowest in children aged 5-14 years (Figure S2b). In this age group, mortality declined from 1.4/1000 PYO in 2003-2006 to 0.8/1000 PYO in 2007-2010 (Table 2) and varied little thereafter. In adults, the steepest period-to-period decline (35%) was seen in the age group 15-54 years between the periods 2003-06 and 2007-10. Mortality changed very little over time for those aged ≥75 years. Overall, mortality was higher in males than females at all ages with differences being greater in adulthood than in childhood.

**Table 2:** Mortality rates per 1000 PYO by age stratum and time period

| Age | 2003-06 | 2007-10 | 2011-14 | 2015-18 |
| --- | --- | --- | --- | --- |
| **0-28 days** |  |  |  |  |
| Total | 330.6 | 200.8 | 174.7 | 172.6 |
| Female | 283.0 | 172.7 | 173.3 | 141.6 |
| Male | 377.2 | 228.5 | 176.1 | 202.8 |
| **29-365 days** |  |  |  |  |
| Total | 18.7 | 11.9 | 10.1 | 8.2 |
| Female | 19.1 | 11.5 | 9.0 | 8.3 |
| Male | 18.4 | 12.2 | 11.2 | 8.0 |
| **<1 year** |  |  |  |  |
| Total | 41.6 | 25.7 | 22.1 | 19.8 |
| Female | 38.4 | 23.3 | 20.9 | 17.8 |
| Male | 44.8 | 28.0 | 23.3 | 21.8 |
| **1-4 years** |  |  |  |  |
| Total | 6.0 | 3.0 | 2.6 | 2.0 |
| Female | 5.9 | 2.7 | 2.5 | 1.7 |
| Male | 6.2 | 3.2 | 2.7 | 2.3 |
| **0-4 years** |  |  |  |  |
| Total | 13.1 | 7.4 | 6.4 | 5.4 |
| Female | 12.4 | 6.7 | 6.0 | 4.7 |
| Male | 13.9 | 8.1 | 6.7 | 6.0 |
| **5-14 years** |  |  |  |  |
| Total | 1.4 | 0.8 | 0.9 | 0.8 |
| Female | 1.2 | 0.7 | 0.8 | 0.7 |
| Male | 1.6 | 0.9 | 1.0 | 1.0 |
| **15-54 years** |  |  |  |  |
| Total | 5.7 | 3.7 | 3.2 | 2.9 |
| Female | 5.6 | 3.6 | 3.0 | 2.7 |
| Male | 5.8 | 3.9 | 3.6 | 3.2 |
| **55-74 years** |  |  |  |  |
| Total | 25.2 | 23.3 | 22.5 | 18.8 |
| Female | 21.1 | 19.3 | 17.5 | 13.6 |
| Male | 30.8 | 28.9 | 29.8 | 26.8 |
| **≥75 years** |  |  |  |  |
| Total | 76.6 | 79.9 | 73.5 | 75.7 |
| Female | 69.5 | 70.4 | 58.0 | 63.0 |
| Male | 84.3 | 91.4 | 94.5 | 96.2 |

### Survival

Median survival increased from 65 in 2003 to 77 in 2018. Females had higher survival throughout the study period and died, on average, 6 years later than males (Figure 1). The survival functions for males and females are similar until approximately 40 years of age after which females experience better survival (Figure 2 A-D). The improvement in survival over time was also greater in females than males (Figure 2 E-F).

**Figure 1.**
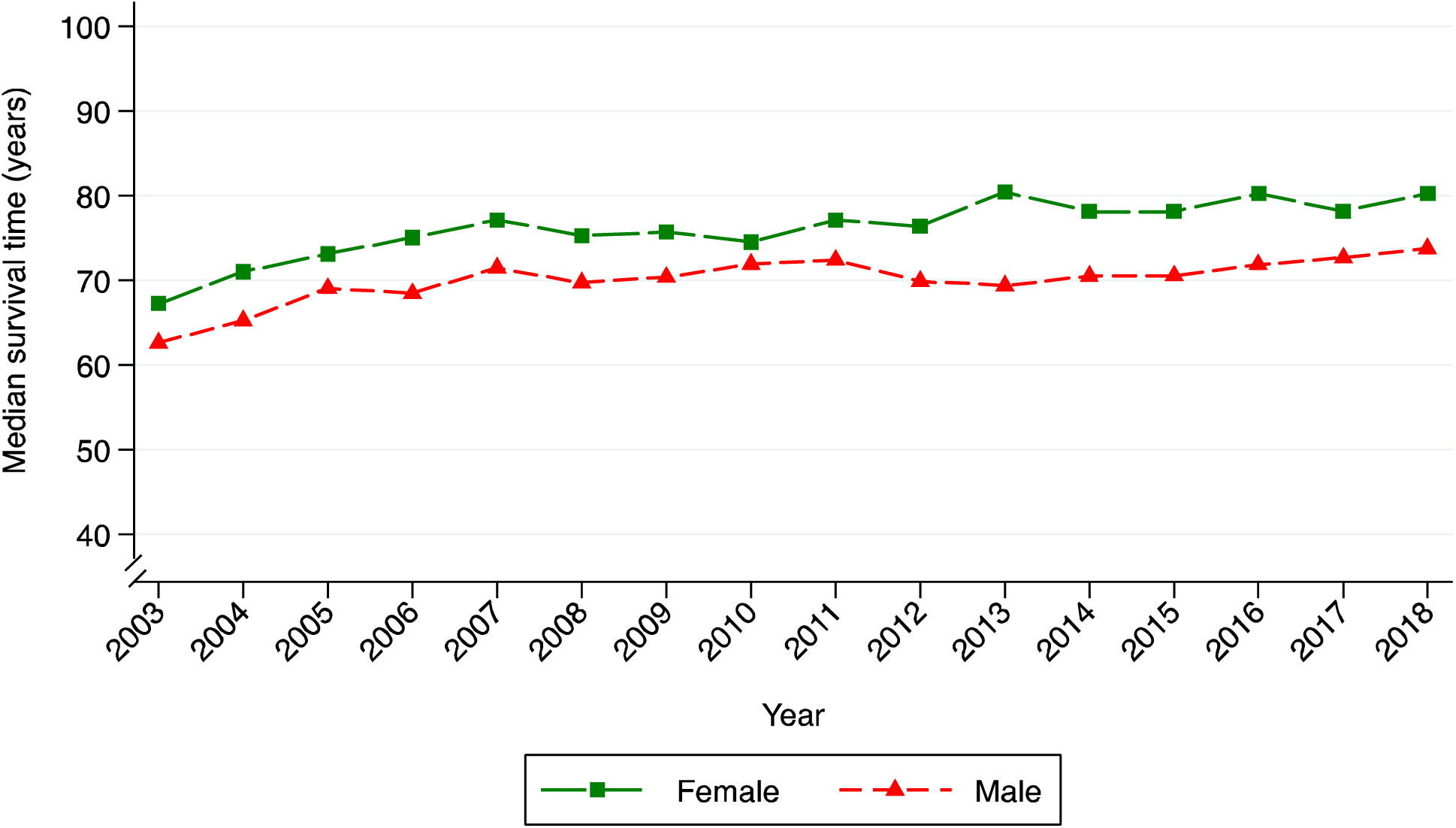
Median age at death based on the Kaplan-Meier method

**Figure 2.**
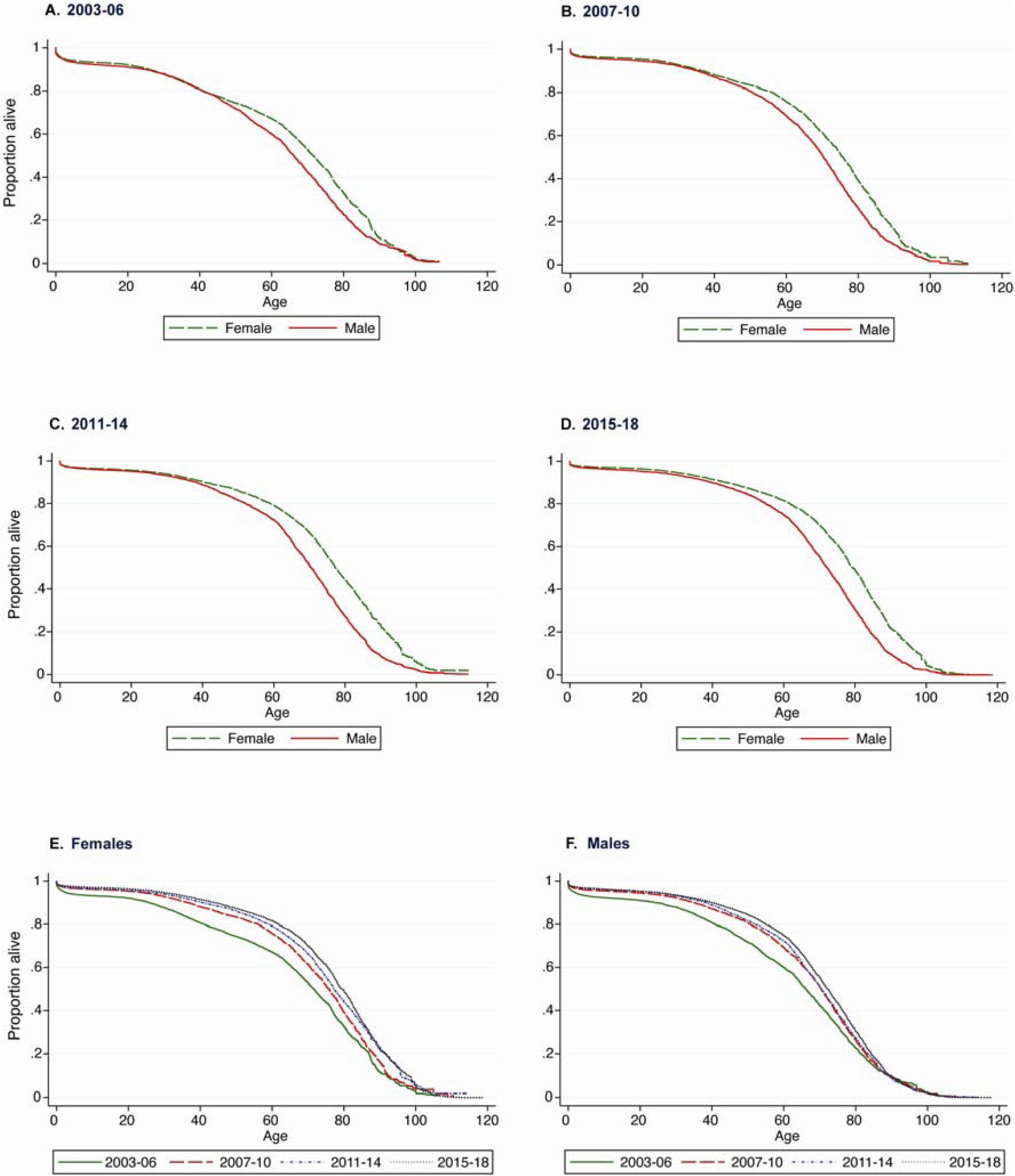
Kaplan-Meier Survival Curves for the KHDSS population by time period and sex

Mean life expectancy, estimated using the abridged life table method, is 4-8 years lower than Kaplan-Meier median survival estimates (Table S3). Tables S4a to S4h show the sex-specific abridged life tables for each of the 4-year periods. The life expectancy estimates calculated using the MADIMAH technique differ little from those estimated using the standard technique (Tables S5a to S5h).

### Seasonality and trend

Figure 3a shows the age-specific mortality rates by calendar month for each of the analysis periods. A seasonal pattern appears only in children aged 1-4 years in the first period. Table S6 shows the estimated high and low mortality months. The months with the highest mortality for neonates, children aged 29-365 days and those aged 1-4 years were February, June, and July respectively. For adults aged 15-74 and ≥75 years, high mortality months were June and August, respectively.

**Figure 3a.**
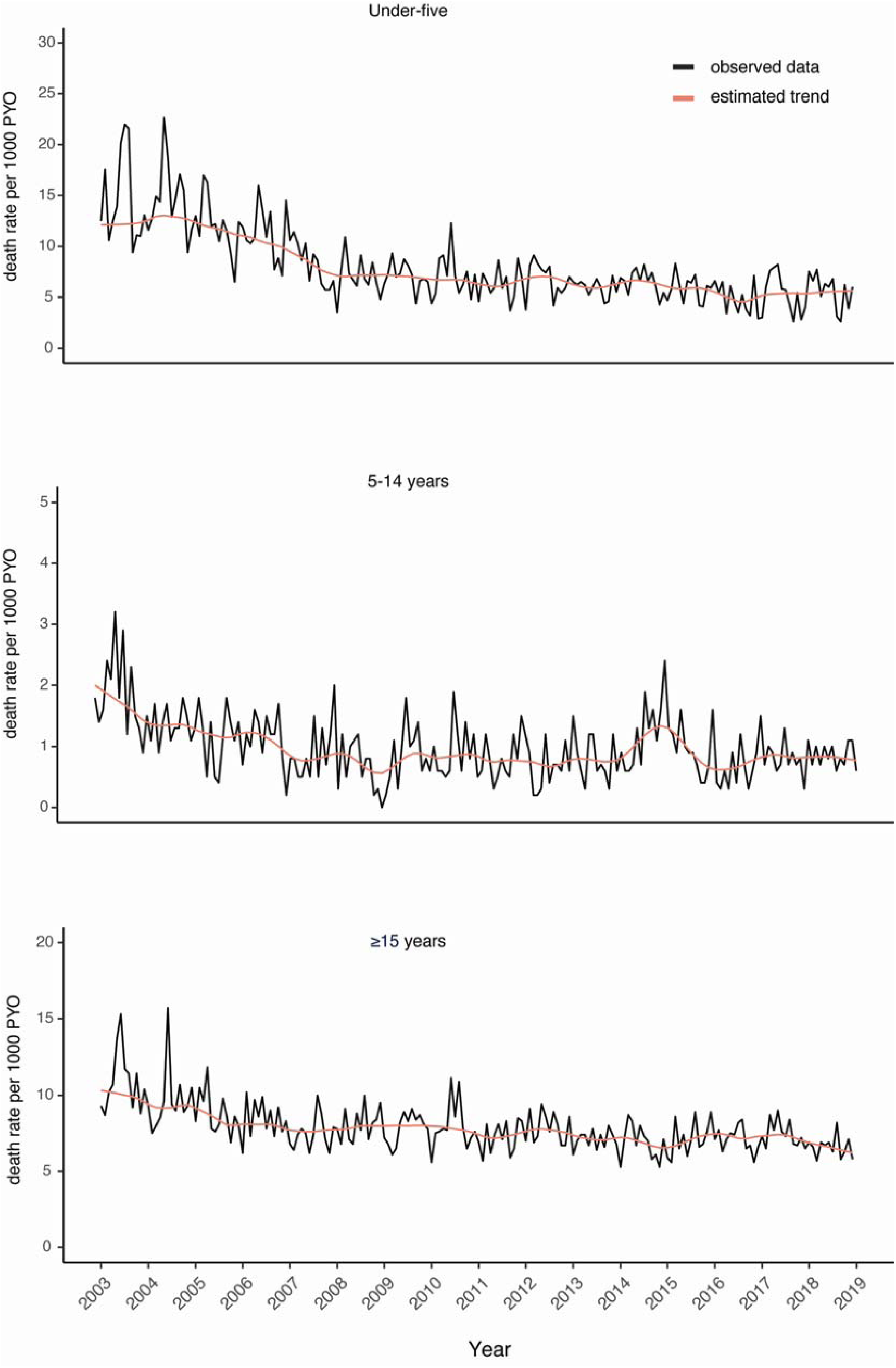
Seasonal mortality rates (by calendar month) by age group and time period

Figure 3b shows the trend in monthly mortality in three broad age groups. For children under five years, there was a steep decline in mortality from 2003 to 2008 followed by a gentle decline in the subsequent years. Within the same period, there was a similar pattern of decline in mortality rates in the older age groups. Figures S3a to S3g show the 16-year mortality trends in finer age strata for both children and adults.

**Figure 3b.**
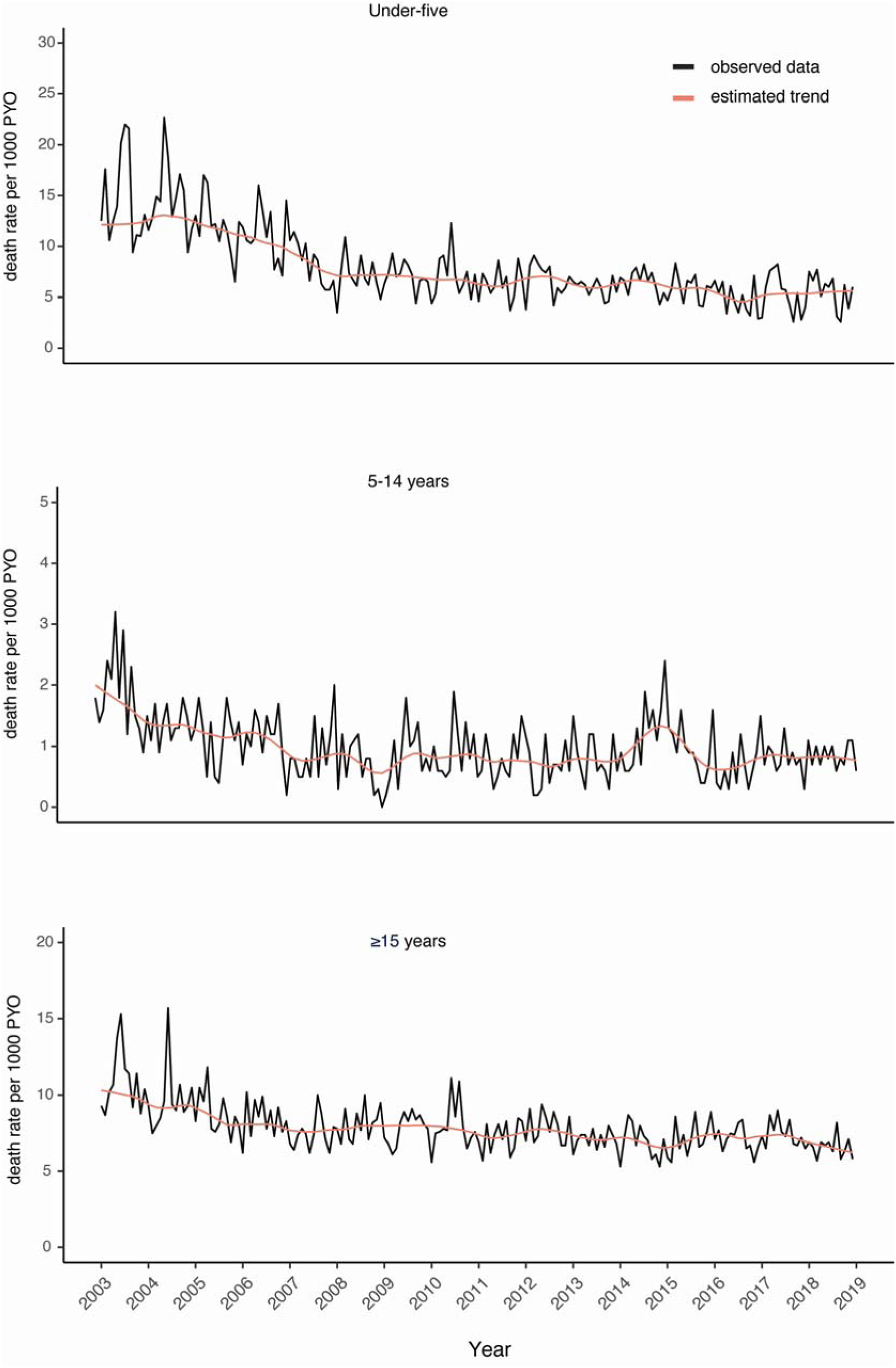
Monthly mortality rates by age over the entire 16-year period

### Geographical heterogeneity in survival and mortality

The largest span in median survival between locations was 7 years (highest in Gede, lowest in Junju and Tezo) in the period 2003-06 (Table S7). The span between highest and lowest location was reduced by 1 year in each of the subsequent 4-year study periods. Gains in life expectancy varied by location with one location (Sokoke) gaining 10 years after the first period and another location (Matsangoni) remaining unchanged. Changes in life expectancy between the periods 2003-06 and 2007-10 were greater in magnitude in comparison to any other two periods.

Figure 4 shows the geographical distribution of the age-standardized mortality rates for each population in the 15 locations over time. Overall mortality declined in all the locations but for those aged ≥55 years the decline was slow in some locations. (Figure S4a & S4b). Table 3 summarises the geographical variation in mortality. After accounting for age and sex, variation was greatest in the period 2003-06 with 95% of the location-specific rates lying within 16% of the median rate in this period. In the periods 2007-10 and 2011-14, we observed a decrease in variation with 95% of the location-specific rates lying within 9% of the median rate in both periods followed by an increase in 2015-18 at 14%. Between-location variation was significant in the 2003-06 and 2015-18 periods. There were no significant differences in geographical variation in mortality between 2003-06 and any of the subsequent periods. The output from the random effects Poisson regression model is summarized in Table S8.

**Figure 4.**
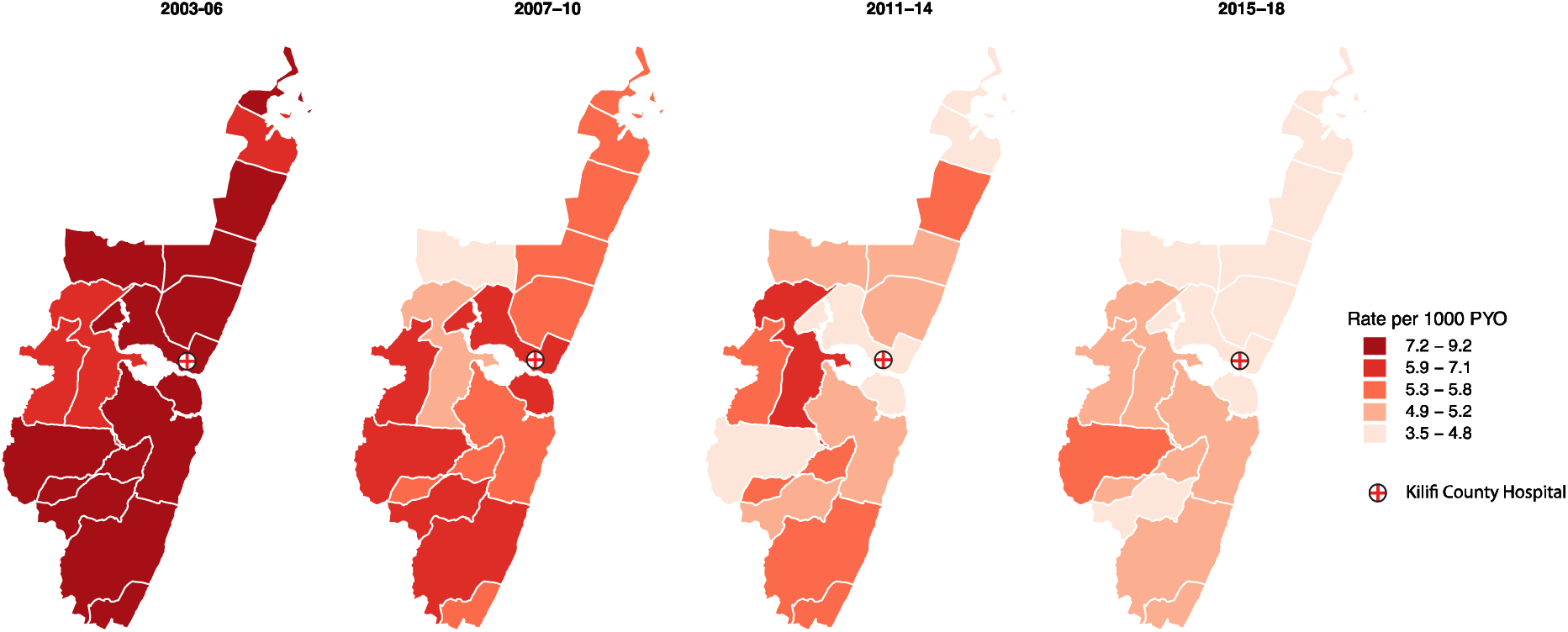
Age-standardised mortality rates by location and time period. Mortality rates were standardised to the KHDSS population structure in 2011

**Table 3.** Variation of location-specific log_e_ mortality rates (per 1000 PYO) adjusted by age and sex.

| Period | SD of log rates<sup>a</sup> | 95% normal approximation<sup>b</sup> | P-value<br>(LR test of between-location variation)<sup>c</sup> | Differences between SDs<sup>d</sup> | P-value<br>(z-test for equality of SD)<sup>e</sup> |
| --- | --- | --- | --- | --- | --- |
| 2003-06 | 0.0778 | 1.16 | <0.001 |  |  |
| 2007-10 | 0.0444 | 1.09 | 0.072 | 0.033 | 0.267 |
| 2011-14 | 0.0433 | 1.09 | 0.067 | 0.034 | 0.246 |
| 2015-18 | 0.0670 | 1.14 | 0.002 | 0.011 | 0.715 |
| 2003-18 | 0.0498 | 1.10 | <0.001 |  |  |

Brief description of the analysis: SD of log rates are derived from a random effect Poisson regression model with administrative location included as the random effect. Geographical variation is quantified via SD of log rates, which is the square root of the variance of the random effect.
<sup>a</sup> Standard deviation - geographical (based on 15 administrative locations) variation in mortality rates. Calculated as the square root of the variance of the random effect.
<sup>b</sup> Approximation of the interval, about the overall median rate, that contains 95% of location-specific rates. Based on the normal distribution assumption that 95% of observations fall within 1.96 standard deviations of the median. Calculated as $\exp(\text{SD of log rates} \times 1.96)$ . An example of the interpretation; in the period 2003-06, 95% of location-specific rates were within 16% of the median rate
<sup>c</sup> Likelihood ratio test comparing the fitted random effects model with that of a conventional Poisson model that does not include the random effect. Null hypothesis: no between-location variation in mortality rates
<sup>d</sup> Difference in SD between 2003-06 and subsequent periods
<sup>e</sup> P-value from test of equality of SDs between 2003-06 and subsequent periods

## DISCUSSION

In the Kilifi Health and Demographic Surveillance System, overall mortality rates declined steeply in the first four years of the study period in all age-sex groups, except in older adults, and then declined much more slowly in the subsequent 12 years. Neonatal and under-five mortality rates declined by 48% and 59%, respectively, between the first period and the last. The mortality reduction observed between the first and subsequent 4-year time periods was greatest among those aged 1-4 years. Median survival was greater in women, by 6 years compared to men, and increased in both sexes by approximately 12 years during the study period. Seasonal effects on mortality were only evident in children aged 1-4 years and only in the first four-year period. Finally, location-specific mortality varied from the median value by +/-10%, which represents an important inequality. There was no evidence that this variation has improved over time.

In LMICs, adult mortality has been characterized by higher rates in females between 15-34 years because of deaths related to childbirth and possibly an earlier age of infection by HIV (29, 30). We observed this phenomenon in the first four years of the study period, but it was later reversed with men being at a greater risk of dying (Figure S5). This pattern is consistent with mortality sex ratios before and after the rapid initiation of HIV care and antiretroviral therapy in LMICs between 2003-2018 with evidence showing that women have benefitted more from the expansion of HIV treatment programs than men (31, 32). In Kilifi, the prevalence of HIV infection among women attending ante-natal at KCH ranged between 3.8%-4.4% between 2005-2009 and 2.0%-3.7% between 2010-2016 with a clear decline from 2010 (5). The phenomenon of women living longer than men is a common observation across populations (33-38) and may be related, in part, to the cardioprotective effect of oestrogen. Following menopause, oestrogen levels decline, and women become susceptible to cardiovascular disease (39). In Kilifi between 2007-12, the incidence of admission to hospital with circulatory system diseases among adults >35 years old was higher in men than women (13) and the mortality fraction due to cardiovascular disease among those aged ≥65 years was marginally higher in men compared to women (20)

Sub-national variation in child mortality, which is driven in part by inequitable distribution of health services and interventions, is a common observation across SSA (40-44). In Kilifi, the magnitude of the overall variation can be understood as meaning that one location (5% of all locations) experienced a mortality rate that lies beyond +/-10% of the average mortality rate. The magnitude of variation did not change significantly over time, which suggests sustained inequitable distribution in public health services and access to health care if we consider geographical variation in mortality as the measure of equity.

The decline in child mortality in Kilifi is consistent with independent observations over an extended period (Table S9). DHS data from Coast Province, which included Kilifi County, showed a reduction in child deaths (<5 years), per 1000 live births, from 116 to 57 between1993-2013. Where the two studies overlap in time (2004-2013), the DHS mortality ratios are higher than the KHDSS estimates; for example, the infant mortality ratios (IMR) are 31 in KHDSS and 44 in the Coastal DHS whilst the under-5 mortality ratios (U5MR) are 46 and 57, respectively. The IMR estimated from the national census in 2009 was 42 per 1000 live births in Kilifi County for the 12 months preceding the census; the equivalent figure from KHDSS in 2008 was 25. For the U5MR the national census and KHDSS estimates were 57 and 38, respectively, per 1000 live births. Both the DHS and census rely on cross-sectional surveys and recall methods for death ascertainment whereas the KHDSS measures mortality directly from a cohort. Paradoxically, the methods that are dependent on potentially unreliable recall provide higher estimates of the number of deaths. The differences between the two methods are more likely to be driven by different definitions of their target populations. DHS and census methods capture all residents observed at one point in time within the geographical locale; some of these may not meet the residence requirement of the HDSS cohort. These requirements include that they are, or intend to be, resident in this household for at least 3 months. Furthermore, the DHS covers the whole of Coast Province, which has 4 counties in addition to Kilifi and, even within Kilifi, the KHDSS is a sub-population (approximately 40%) of the county. The overall numbers of child mortality from the HDSS, DHS and Census datasets may be different but the trends are consistent with each other and are also consistent with an analysis of multiple disparate datasets that shows a decline in child mortality beginning as far back as 1965 (40).

This descriptive analysis lays out the baseline trends in mortality in Kilifi over time. Several additional data sources may help to explore the underlying causes of these trends and geographic patterns. Within KHDSS there are data on the changing morbidity experience of residents from hospital records over the same period; for example, the incidence of admission to hospital with malaria declined sharply between 2003-2006 (45). Similar declines in LMICs have largely been attributed to reductions in malaria transmission following a high coverage of control measures (46). The declining trend in malaria admissions in Kilifi is similar to the all-cause child mortality trend we have observed. Previous studies have reported a substantial indirect contribution of malaria to all-cause child mortality (47-49). A study in this population found that malaria infection strongly predisposes individuals to bacteremia, a major cause of childhood death, and could account for at least 50% of all bacteremia cases in children (50). The geographical heterogeneity in mortality (Figure 4) appears similar to the geographical heterogeneity in malaria (51), as has been observed with national heterogeneity in malaria and mortality (52). In Kilifi, we observed a significant reduction in childhood morbidity after the introduction of *Haemophilus influenzae* type b conjugate vaccine in 2001 (53), 10-valent pneumococcal conjugate vaccine in 2011 (5, 15) and rotavirus vaccine in 2014 (16) but their effects on mortality have not yet been established. Finally, verbal autopsy data are also available to explore changes in causes of death over time (20). The integration of these sources of data is beyond the scope of the present analysis.

Between 2003 and 2018, Kenya has experienced average GDP growth of 5.24% per annum and a population growth of 2.63% per annum (54). The area of KHDSS is typical of much of sub-Saharan Africa, being largely rural with a central town of approximately 43,000 residents (14% of the KHDSS). The detailed surveillance reported here over 16 years in KHDSS illustrates a clear improvement in mortality rates at all ages below 75 years but the pace of improvement has declined markedly over time and the geographical distribution of mortality rates is not homogeneous. These findings highlight opportunities for intervention and improvement across a wide range of health, social and economic domains.

## Data Availability

Data that support the findings of this study are available from the KEMRI Institutional Data Access/Ethics Committee, for researchers who meet the criteria for access to confidential data. Details of the criteria can be found in the KEMRI-Wellcome data sharing guidelines. Access to data is provided via the KEMRI-Wellcome Data Governance Committee.

## Data Availability

Data that support the findings of this study are available from the KEMRI Institutional Data Access/Ethics Committee, for researchers who meet the criteria for access to confidential data. Details of the criteria can be found in the KEMRI-Wellcome <u>data sharing guidelines</u>. Access to data is provided via the KEMRI-Wellcome Data Governance Committee.

## Abbreviations

LMICs: Low- and Middle-Income Countries
sSA: sub-Saharan Africa
CRVS: Civil Registration and Vital Statistics Systems
DHS: Demographic and Health Surveys
HDSSs: Health and Demographic Surveillance Systems
KWTRP: KEMRI-Wellcome Trust Research Programme
KHDSS: Kilifi Health and Demographic Surveillance System
KCH: Kilifi County Hospital
INDEPTH: International Network for the Demographic Evaluation of Populations and Their Health
MySQL: Structured query language
PYO: Person years of observation
KM: Kaplan-Meier
MADIMAH: Multi-centre Analysis of the Dynamics of Internal Migration and Health
STL: Seasonal and Trend using LOESS
HIV: Human immunodeficiency virus
IMR: Infant mortality rate
U5MR: Under-five mortality rate
GDP: Gross domestic product

## Contributions

Conceptualization: JAGS, KM, TNW and EB. Data collection and preparation: MO, EB, AN, DA, CN, ET, DW, MW, NK, MK, GN. Analysis: MO, CB, JAGS Interpretation: MO, JAGS, CB, AN, AD, JM, AG, BT, PB, KM, JM, EB, TNW. Drafting the manuscript: MO, JAGS, CB, PB. Resources and funding acquisition: PB, JAGS, TNW, KM, BT, AN. All authors critically reviewed the article and approved the final version for submission.

## Acknowledgements

We gratefully thank the residents of Kilifi who have participated in the surveillance activities of the KHDSS. We acknowledge the tremendous work of the census field staff and data supervisors who collect and process this information and the Community Liaison Group who run the community engagement programmes. We would particularly like to acknowledge the important contributions of Victoria Nyaga, John Ojal and Hellen Gatakaa (former HDSS statisticians). This article is published with the permission of the Director of the Kenya Medical Research Institute.

