## Supplementary Information for "Mortality in Rural Coastal Kenya measured using the Kilifi Health and Demographic Surveillance System: A 16-year descriptive analysis"

Authors and Affiliations:

Mark Otiende^1*^, Evasius Bauni^1^, Amek Nyaguara^1^, David Amadi^1^, Christopher Nyundo^1^, Emmanuel Tsory^1^, David Walumbe^1^, Michael Kinuthia^1^, Norbert Kihuha^1^, Michael Kahindi^1^, Gideon Nyutu^1^, Jennifer Moisi^1^, Amare Deribew^1^, Ambrose Agweyu^1^, Kevin Marsh^3^, Benjamin Tsofa^1^, Philip Bejon^1,3^, Christian Bottomley^2^, Thomas N. Williams^1^, J. Anthony G. Scott ^1,2^

1. KEMRI-Wellcome Trust Research Programme, Kilifi, Kenya
2. Department of Infectious Disease Epidemiology, London School of Hygiene & Tropical Medicine, London, UK
3. Nuffield Department of Clinical Medicine, University of Oxford, Oxford, UK

* Corresponding Author

Table S1. Quality of age and sex variables

| Year | Newly registered residents | Whipple's Index | Sex ratio |
| --- | --- | --- | --- |
| 2003 | 46,557 | 102.1 | 89:100 |
| 2004 | 26,616 | 107.6 | 89:100 |
| 2005 | 24,274 | 123.2 | 90:100 |
| 2006 | 23,409 | 123.2 | 92:100 |
| 2007 | 20,419 | 89.5 | 93:100 |
| 2008 | 24,214 | 104.0 | 98:100 |
| 2009 | 29,023 | 111.2 | 97:100 |
| 2010 | 25,620 | 124.2 | 96:100 |
| 2011 | 20,165 | 104.6 | 96:100 |
| 2012 | 24,465 | 92.7 | 97:100 |
| 2013 | 18,744 | 106.9 | 95:100 |
| 2014 | 22,002 | 100.2 | 95:100 |
| 2015 | 17,324 | 110.2 | 96:100 |
| 2016 | 17,167 | 98.1 | 91:100 |
| 2017 | 14,263 | 93.9 | 91:100 |
| 2018 | 17,749 | 105.2 | 96:100 |

- An index that is <105 indicates that there is no preference for reporting ages ending in 0 and 5.

Whipple’s index and sex ratio was calculated for 2003 based on the age and sex reported in the baseline census done in 2000-1. For all subsequent years the indices and the ratios are based on the declared age of all in-migrants.

- Whipple’s Index and sex ratio calculated based on age and sex reported at first contact

Table S2. Mortality ratios expressed per 1000 live births

| **Age** | **2003-06** | **2007-10** | **2011-14** | **2015-18** |
| --- | --- | --- | --- | --- |
| **0-28 days** |  |  |  |  |
| Total | 25.0 | 15.3 | 13.2 | 13.1 |
| Female | 21.5 | 13.2 | 13.2 | 10.7 |
| Male | 28.5 | 17.4 | 13.3 | 15.3 |
| **29-365 days** |  |  |  |  |
| Total | 17.9 | 11.5 | 9.8 | 8.1 |
| Female | 18.4 | 11.1 | 8.8 | 8.3 |
| Male | 17.5 | 11.8 | 10.8 | 8.0 |
| **<1 year** |  |  |  |  |
| Total | 42.9 | 26.8 | 23.0 | 21.2 |
| Female | 39.9 | 24.3 | 21.9 | 19.0 |
| Male | 46.0 | 29.1 | 24.1 | 23.3 |
| **1-4 years** |  |  |  |  |
| Total | 25.0 | 12.8 | 11.3 | 9.1 |
| Female | 24.5 | 11.6 | 10.8 | 7.5 |
| Male | 25.4 | 13.9 | 11.8 | 10.5 |
| **U5 years** |  |  |  |  |
| Total | 67.9 | 39.5 | 34.3 | 30.3 |
| Female | 64.4 | 35.9 | 32.7 | 26.5 |
| Male | 71.3 | 43.0 | 35.9 | 33.9 |

Table S3. Comparison of mean life expectancy at birth based on abridged lifetable method and median age at death according to the KM method

| **Method** | **Sex** | **2003-06** | **2007-10** | **2011-14** | **2015-18** |
| --- | --- | --- | --- | --- | --- |
| Median age at death (KM) | Female | 72 | 76 | 77 | 79 |
|  | Male | 66 | 70 | 70 | 72 |
| Mean life expectancy (abridged life table method) | Female | 64 | 68 | 72 | 74 |
|  | Male | 60 | 65 | 66 | 68 |

Table S4a. Lifetable for the period 2003-06 for females (conventional method)

| Age (years) | nDx | nPYx | nMx | SE nMx | nqx | SE nqx | lx | SE lx | ndx | nLx | Tx | ex (years) | SE ex (years) |
| --- | --- | --- | --- | --- | --- | --- | --- | --- | --- | --- | --- | --- | --- |
| <1 | 601 | 15663.74 | 0.0384 | 0.0016 | 0.0386 | 0.0015 | 100000 |  | 3860 | 97044 | 6396268 | **64** | 0.3349 |
| 1-4 | 370 | 62813.97 | 0.0059 | 0.0003 | 0.0231 | 0.0012 | 96140 | 154 | 2224 | 378710 | 6299224 | **66** | 0.3317 |
| 5-9 | 101 | 70592.18 | 0.0014 | 0.0001 | 0.0070 | 0.0007 | 93916 | 189 | 661 | 467620 | 5920514 | **63** | 0.3300 |
| 10-14 | 53 | 59564.64 | 0.0009 | 0.0001 | 0.0045 | 0.0006 | 93255 | 199 | 417 | 465244 | 5452894 | **58** | 0.3295 |
| 15-19 | 73 | 46774.66 | 0.0016 | 0.0002 | 0.0079 | 0.0009 | 92838 | 206 | 736 | 462518 | 4987650 | **54** | 0.3292 |
| 20-24 | 124 | 33661.29 | 0.0037 | 0.0003 | 0.0184 | 0.0016 | 92102 | 222 | 1692 | 456461 | 4525132 | **49** | 0.3284 |
| 25-29 | 201 | 32937.36 | 0.0061 | 0.0004 | 0.0299 | 0.0021 | 90410 | 265 | 2707 | 445145 | 4068671 | **45** | 0.3251 |
| 30-34 | 202 | 25503.89 | 0.0079 | 0.0006 | 0.0386 | 0.0027 | 87703 | 319 | 3386 | 430209 | 3623526 | **41** | 0.3216 |
| 35-39 | 176 | 20094.67 | 0.0088 | 0.0007 | 0.0427 | 0.0031 | 84317 | 385 | 3599 | 411485 | 3193317 | **38** | 0.3153 |
| 40-44 | 139 | 17872.24 | 0.0078 | 0.0007 | 0.0388 | 0.0032 | 80718 | 458 | 3132 | 395914 | 2781832 | **34** | 0.3057 |
| 45-49 | 145 | 17459.10 | 0.0083 | 0.0007 | 0.0405 | 0.0033 | 77586 | 512 | 3140 | 379243 | 2385918 | **31** | 0.2979 |
| 50-54 | 104 | 12129.74 | 0.0086 | 0.0008 | 0.0419 | 0.0040 | 74446 | 555 | 3119 | 364669 | 2006675 | **27** | 0.2933 |
| 55-59 | 127 | 10458.17 | 0.0121 | 0.0011 | 0.0580 | 0.0050 | 71327 | 611 | 4136 | 345403 | 1642006 | **23** | 0.2868 |
| 60-64 | 162 | 8462.83 | 0.0191 | 0.0015 | 0.0912 | 0.0068 | 67191 | 678 | 6130 | 322477 | 1296603 | **19** | 0.2813 |
| 65-69 | 161 | 5743.02 | 0.0280 | 0.0022 | 0.1328 | 0.0097 | 61061 | 770 | 8109 | 282733 | 974126 | **16** | 0.2778 |
| 70-74 | 158 | 4149.32 | 0.0381 | 0.0030 | 0.1729 | 0.0125 | 52952 | 908 | 9155 | 242126 | 691393 | **13** | 0.2663 |
| 75-79 | 132 | 2334.19 | 0.0566 | 0.0049 | 0.2558 | 0.0192 | 43797 | 1005 | 11202 | 186073 | 449267 | **10** | 0.2580 |
| 80-84 | 86 | 1259.76 | 0.0683 | 0.0074 | 0.2892 | 0.0263 | 32595 | 1182 | 9425 | 141346 | 263194 | **8** | 0.1986 |
| 85+ | 89 | 820.71 | 0.1084 | 0.0116 | 1.0000 | 0.0000 | 23170 | 1205 | 23170 | 121848 | 121848 | **5** | 0.0000 |

Table S4b. Lifetable for the period 2003-06 for males (conventional method)

| Age (years) | nDx | nPYx | nMx | SE nMx | nqx | SE nqx | lx | SE lx | ndx | nLx | Tx | ex (years) | SE ex (years) |
| --- | --- | --- | --- | --- | --- | --- | --- | --- | --- | --- | --- | --- | --- |
| <1 | 712 | 15905.91 | 0.0448 | 0.0017 | 0.0449 | 0.0016 | 100000 |  | 4485 | 96370 | 6029976 | **60** | 0.3521 |
| 1-4 | 393 | 63763.68 | 0.0062 | 0.0003 | 0.0243 | 0.0012 | 95515 | 164 | 2317 | 376328 | 5933606 | **62** | 0.3524 |
| 5-9 | 141 | 71170.58 | 0.0020 | 0.0002 | 0.0097 | 0.0008 | 93198 | 197 | 907 | 463318 | 5557278 | **60** | 0.3528 |
| 10-14 | 75 | 62042.21 | 0.0012 | 0.0001 | 0.0060 | 0.0007 | 92291 | 209 | 554 | 459929 | 5093960 | **55** | 0.3531 |
| 15-19 | 82 | 48364.59 | 0.0017 | 0.0002 | 0.0087 | 0.0010 | 91737 | 218 | 798 | 456679 | 4634031 | **51** | 0.3533 |
| 20-24 | 61 | 26411.29 | 0.0023 | 0.0003 | 0.0118 | 0.0015 | 90939 | 233 | 1071 | 452037 | 4177352 | **46** | 0.3533 |
| 25-29 | 95 | 21553.68 | 0.0044 | 0.0005 | 0.0217 | 0.0022 | 89868 | 269 | 1949 | 444104 | 3725315 | **41** | 0.3512 |
| 30-34 | 124 | 17851.71 | 0.0069 | 0.0006 | 0.0341 | 0.0030 | 87919 | 329 | 2996 | 431843 | 3281211 | **37** | 0.3475 |
| 35-39 | 143 | 13867.82 | 0.0103 | 0.0009 | 0.0500 | 0.0041 | 84923 | 414 | 4245 | 413088 | 2849368 | **34** | 0.3418 |
| 40-44 | 113 | 11999.63 | 0.0094 | 0.0009 | 0.0458 | 0.0042 | 80678 | 526 | 3698 | 394203 | 2436280 | **30** | 0.3309 |
| 45-49 | 145 | 9975.65 | 0.0145 | 0.0012 | 0.0694 | 0.0056 | 76980 | 606 | 5341 | 370706 | 2042077 | **27** | 0.3223 |
| 50-54 | 162 | 9215.90 | 0.0176 | 0.0014 | 0.0846 | 0.0064 | 71639 | 709 | 6059 | 344994 | 1671371 | **23** | 0.3096 |
| 55-59 | 134 | 7412.65 | 0.0181 | 0.0016 | 0.0864 | 0.0071 | 65580 | 793 | 5668 | 313775 | 1326377 | **20** | 0.3002 |
| 60-64 | 170 | 6026.09 | 0.0282 | 0.0022 | 0.1319 | 0.0094 | 59912 | 864 | 7902 | 282699 | 1012602 | **17** | 0.2916 |
| 65-69 | 173 | 4217.89 | 0.0410 | 0.0031 | 0.1877 | 0.0129 | 52010 | 940 | 9760 | 235184 | 729903 | **14** | 0.2864 |
| 70-74 | 169 | 3336.67 | 0.0506 | 0.0039 | 0.2226 | 0.0151 | 42250 | 1020 | 9404 | 189780 | 494719 | **12** | 0.2709 |
| 75-79 | 151 | 2029.57 | 0.0744 | 0.0061 | 0.3129 | 0.0211 | 32846 | 1018 | 10276 | 134285 | 304939 | **9** | 0.2664 |
| 80-84 | 105 | 1181.01 | 0.0889 | 0.0087 | 0.3573 | 0.0280 | 22570 | 1010 | 8064 | 93371 | 170654 | **8** | 0.2164 |
| 85+ | 84 | 823.72 | 0.1020 | 0.0112 | 1.0000 | 0.0000 | 14506 | 903 | 14506 | 77283 | 77283 | **5** | 0.0000 |

Table S4c. Lifetable for the period 2007-10 for females (conventional method)

| Age (years) | nDx | nPYx | nMx | SE nMx | nqx | SE nqx | lx | SE lx | ndx | nLx | Tx | ex (years) | SE ex (years) |
| --- | --- | --- | --- | --- | --- | --- | --- | --- | --- | --- | --- | --- | --- |
| <1 | 384 | 16465.00 | 0.0233 | 0.0012 | 0.0237 | 0.0012 | 100000 |  | 2370 | 98149 | 6936354 | **69** | 0.3061 |
| 1-4 | 183 | 68628.12 | 0.0027 | 0.0002 | 0.0105 | 0.0008 | 97630 | 119 | 1027 | 387931 | 6838205 | **70** | 0.3013 |
| 5-9 | 65 | 78421.24 | 0.0008 | 0.0001 | 0.0041 | 0.0005 | 96603 | 140 | 398 | 481997 | 6450274 | **67** | 0.2997 |
| 10-14 | 45 | 68604.65 | 0.0007 | 0.0001 | 0.0033 | 0.0005 | 96205 | 148 | 317 | 480251 | 5968277 | **62** | 0.2991 |
| 15-19 | 55 | 51468.15 | 0.0011 | 0.0001 | 0.0054 | 0.0007 | 95888 | 155 | 517 | 478153 | 5488026 | **57** | 0.2986 |
| 20-24 | 81 | 40458.32 | 0.0020 | 0.0002 | 0.0101 | 0.0011 | 95371 | 169 | 968 | 474691 | 5009873 | **53** | 0.2975 |
| 25-29 | 108 | 31277.67 | 0.0035 | 0.0003 | 0.0171 | 0.0016 | 94403 | 199 | 1619 | 468582 | 4535182 | **48** | 0.2951 |
| 30-34 | 140 | 30683.66 | 0.0046 | 0.0004 | 0.0228 | 0.0019 | 92784 | 249 | 2117 | 458654 | 4066600 | **44** | 0.2904 |
| 35-39 | 137 | 23987.05 | 0.0057 | 0.0005 | 0.0281 | 0.0024 | 90667 | 301 | 2552 | 447490 | 3607946 | **40** | 0.2855 |
| 40-44 | 94 | 17438.30 | 0.0054 | 0.0006 | 0.0269 | 0.0027 | 88115 | 363 | 2373 | 434163 | 3160456 | **36** | 0.2786 |
| 45-49 | 103 | 18675.85 | 0.0055 | 0.0005 | 0.0277 | 0.0027 | 85742 | 430 | 2378 | 422624 | 2726293 | **32** | 0.2694 |
| 50-54 | 105 | 15614.43 | 0.0067 | 0.0007 | 0.0333 | 0.0032 | 83364 | 479 | 2778 | 409582 | 2303669 | **28** | 0.2641 |
| 55-59 | 145 | 11829.56 | 0.0123 | 0.0010 | 0.0593 | 0.0048 | 80586 | 535 | 4777 | 392821 | 1894087 | **24** | 0.2592 |
| 60-64 | 148 | 9572.35 | 0.0155 | 0.0013 | 0.0744 | 0.0059 | 75809 | 634 | 5641 | 364433 | 1501266 | **20** | 0.2521 |
| 65-69 | 204 | 7782.94 | 0.0262 | 0.0018 | 0.1226 | 0.0080 | 70168 | 738 | 8605 | 328985 | 1136833 | **16** | 0.2447 |
| 70-74 | 151 | 4350.86 | 0.0347 | 0.0028 | 0.1596 | 0.0119 | 61563 | 862 | 9826 | 280939 | 807848 | **13** | 0.2392 |
| 75-79 | 188 | 3435.07 | 0.0547 | 0.0040 | 0.2384 | 0.0152 | 51737 | 1042 | 12332 | 229916 | 526909 | **10** | 0.2179 |
| 80-84 | 100 | 1339.45 | 0.0747 | 0.0075 | 0.3207 | 0.0264 | 39405 | 1120 | 12638 | 161580 | 296993 | **8** | 0.2079 |
| 85+ | 138 | 1273.41 | 0.1084 | 0.0093 | 1.0000 | 0.0000 | 26767 | 1347 | 26767 | 135413 | 135413 | **5** | 0.0000 |

Table S4d. Lifetable for the period 2007-10 for males (conventional method)

| Age (years) | nDx | nPYx | nMx | SE nMx | nqx | SE nqx | lx | SE lx | ndx | nLx | Tx | ex (years) | SE ex (years) |
| --- | --- | --- | --- | --- | --- | --- | --- | --- | --- | --- | --- | --- | --- |
| <1 | 470 | 16772.64 | 0.0280 | 0.0013 | 0.0285 | 0.0013 | 100000 |  | 2847 | 97699 | 6525792 | **65** | 0.3326 |
| 1-4 | 224 | 69241.99 | 0.0032 | 0.0002 | 0.0127 | 0.0008 | 97153 | 129 | 1238 | 385527 | 6428093 | **66** | 0.3305 |
| 5-9 | 79 | 79451.53 | 0.0010 | 0.0001 | 0.0050 | 0.0006 | 95915 | 151 | 475 | 478327 | 6042566 | **63** | 0.3300 |
| 10-14 | 59 | 69833.55 | 0.0008 | 0.0001 | 0.0042 | 0.0005 | 95440 | 160 | 399 | 476083 | 5564239 | **58** | 0.3299 |
| 15-19 | 64 | 53675.66 | 0.0012 | 0.0002 | 0.0060 | 0.0008 | 95041 | 167 | 574 | 473804 | 5088156 | **54** | 0.3299 |
| 20-24 | 58 | 31915.71 | 0.0018 | 0.0002 | 0.0093 | 0.0012 | 94467 | 181 | 875 | 470105 | 4614352 | **49** | 0.3296 |
| 25-29 | 67 | 21517.15 | 0.0031 | 0.0004 | 0.0153 | 0.0019 | 93592 | 214 | 1433 | 464871 | 4144247 | **44** | 0.3277 |
| 30-34 | 95 | 20171.41 | 0.0047 | 0.0005 | 0.0234 | 0.0024 | 92159 | 273 | 2156 | 455289 | 3679376 | **40** | 0.3232 |
| 35-39 | 108 | 16640.92 | 0.0065 | 0.0006 | 0.0321 | 0.0030 | 90003 | 345 | 2885 | 443164 | 3224087 | **36** | 0.3176 |
| 40-44 | 88 | 12830.19 | 0.0069 | 0.0007 | 0.0338 | 0.0035 | 87118 | 432 | 2945 | 427885 | 2780923 | **32** | 0.3100 |
| 45-49 | 98 | 11480.75 | 0.0085 | 0.0009 | 0.0421 | 0.0042 | 84173 | 520 | 3540 | 412399 | 2353038 | **28** | 0.3006 |
| 50-54 | 116 | 9279.31 | 0.0125 | 0.0012 | 0.0606 | 0.0055 | 80633 | 609 | 4884 | 391077 | 1940639 | **24** | 0.2920 |
| 55-59 | 161 | 9021.08 | 0.0178 | 0.0014 | 0.0849 | 0.0064 | 75749 | 722 | 6432 | 363364 | 1549562 | **20** | 0.2809 |
| 60-64 | 166 | 6631.59 | 0.0250 | 0.0020 | 0.1176 | 0.0086 | 69317 | 820 | 8150 | 327087 | 1186198 | **17** | 0.2750 |
| 65-69 | 187 | 5165.85 | 0.0362 | 0.0027 | 0.1659 | 0.0111 | 61167 | 937 | 10147 | 282369 | 859111 | **14** | 0.2677 |
| 70-74 | 180 | 3175.13 | 0.0567 | 0.0042 | 0.2469 | 0.0160 | 51020 | 1036 | 12599 | 221665 | 576742 | **11** | 0.2651 |
| 75-79 | 200 | 2653.02 | 0.0754 | 0.0053 | 0.3139 | 0.0184 | 38421 | 1134 | 12059 | 161919 | 355077 | **9** | 0.2454 |
| 80-84 | 115 | 1180.17 | 0.0974 | 0.0091 | 0.3787 | 0.0278 | 26362 | 1050 | 9983 | 101116 | 193158 | **7** | 0.2420 |
| 85+ | 140 | 1147.45 | 0.1220 | 0.0104 | 1.0000 | 0.0000 | 16379 | 1003 | 16379 | 92042 | 92042 | **6** | 0.0000 |

Table S4e. Lifetable for the period 2011-14 for females (conventional method)

| Age (years) | nDx | nPYx | nMx | SE nMx | nqx | SE nqx | lx | SE lx | ndx | nLx | Tx | ex (years) | SE ex (years) |
| --- | --- | --- | --- | --- | --- | --- | --- | --- | --- | --- | --- | --- | --- |
| <1 | 355 | 16957.35 | 0.0209 | 0.0011 | 0.0215 | 0.0011 | 100000 |  | 2147 | 98236 | 7206812 | **72** | 0.2991 |
| 1-4 | 174 | 70801.24 | 0.0025 | 0.0002 | 0.0098 | 0.0007 | 97853 | 112 | 962 | 389029 | 7108576 | **73** | 0.2938 |
| 5-9 | 80 | 84430.51 | 0.0009 | 0.0001 | 0.0047 | 0.0005 | 96891 | 133 | 454 | 483146 | 6719547 | **69** | 0.2918 |
| 10-14 | 42 | 75651.78 | 0.0006 | 0.0001 | 0.0028 | 0.0004 | 96437 | 141 | 267 | 481458 | 6236401 | **65** | 0.2910 |
| 15-19 | 57 | 56622.87 | 0.0010 | 0.0001 | 0.0050 | 0.0007 | 96170 | 147 | 484 | 479585 | 5754943 | **60** | 0.2906 |
| 20-24 | 63 | 42692.69 | 0.0015 | 0.0002 | 0.0075 | 0.0009 | 95686 | 160 | 713 | 476880 | 5275358 | **55** | 0.2895 |
| 25-29 | 80 | 36249.90 | 0.0022 | 0.0002 | 0.0110 | 0.0012 | 94973 | 182 | 1047 | 472241 | 4798478 | **51** | 0.2874 |
| 30-34 | 116 | 30627.58 | 0.0038 | 0.0004 | 0.0185 | 0.0017 | 93926 | 215 | 1739 | 465709 | 4326237 | **46** | 0.2843 |
| 35-39 | 113 | 28124.82 | 0.0040 | 0.0004 | 0.0200 | 0.0019 | 92187 | 265 | 1845 | 456191 | 3860528 | **42** | 0.2793 |
| 40-44 | 100 | 22827.35 | 0.0044 | 0.0004 | 0.0217 | 0.0021 | 90342 | 312 | 1960 | 446541 | 3404337 | **38** | 0.2746 |
| 45-49 | 79 | 15583.23 | 0.0051 | 0.0006 | 0.0253 | 0.0028 | 88382 | 361 | 2232 | 436747 | 2957796 | **33** | 0.2692 |
| 50-54 | 135 | 19135.41 | 0.0071 | 0.0006 | 0.0352 | 0.0030 | 86150 | 431 | 3030 | 423147 | 2521049 | **29** | 0.2608 |
| 55-59 | 135 | 13813.24 | 0.0098 | 0.0008 | 0.0477 | 0.0040 | 83120 | 490 | 3964 | 405000 | 2097902 | **25** | 0.2565 |
| 60-64 | 164 | 11100.77 | 0.0148 | 0.0012 | 0.0714 | 0.0054 | 79156 | 575 | 5655 | 382647 | 1692902 | **21** | 0.2496 |
| 65-69 | 163 | 8140.16 | 0.0200 | 0.0016 | 0.0968 | 0.0072 | 73501 | 684 | 7114 | 350341 | 1310255 | **18** | 0.2422 |
| 70-74 | 230 | 6512.91 | 0.0353 | 0.0023 | 0.1627 | 0.0098 | 66387 | 817 | 10800 | 305560 | 959914 | **14** | 0.2319 |
| 75-79 | 145 | 3105.15 | 0.0467 | 0.0039 | 0.2102 | 0.0155 | 55587 | 954 | 11685 | 247241 | 654354 | **12** | 0.2212 |
| 80-84 | 137 | 2602.79 | 0.0526 | 0.0045 | 0.2331 | 0.0174 | 43902 | 1152 | 10235 | 193830 | 407113 | **9** | 0.1543 |
| 85+ | 132 | 1431.90 | 0.0922 | 0.0081 | 1.0000 | 0.0000 | 33667 | 1195 | 33667 | 213283 | 213283 | **6** | 0.0000 |

Table S4f. Lifetable for the period 2011-14 for males (conventional method)

| Age (years) | nDx | nPYx | nMx | SE nMx | nqx | SE nqx | lx | SE lx | ndx | nLx | Tx | ex (years) | SE ex (years) |
| --- | --- | --- | --- | --- | --- | --- | --- | --- | --- | --- | --- | --- | --- |
| <1 | 405 | 17410.92 | 0.0233 | 0.0012 | 0.0237 | 0.0012 | 100000 |  | 2366 | 98072 | 6630181 | **66** | 0.3156 |
| 1-4 | 198 | 72231.33 | 0.0027 | 0.0002 | 0.0109 | 0.0008 | 97634 | 116 | 1069 | 387890 | 6532109 | **67** | 0.3131 |
| 5-9 | 97 | 85879.62 | 0.0011 | 0.0001 | 0.0056 | 0.0006 | 96565 | 137 | 542 | 481402 | 6144219 | **64** | 0.3123 |
| 10-14 | 62 | 77436.48 | 0.0008 | 0.0001 | 0.0040 | 0.0005 | 96023 | 147 | 384 | 479109 | 5662817 | **59** | 0.3121 |
| 15-19 | 50 | 58325.17 | 0.0009 | 0.0001 | 0.0045 | 0.0006 | 95639 | 154 | 428 | 477254 | 5183708 | **54** | 0.3120 |
| 20-24 | 55 | 36131.81 | 0.0015 | 0.0002 | 0.0077 | 0.0010 | 95211 | 165 | 735 | 474213 | 4706454 | **49** | 0.3117 |
| 25-29 | 75 | 25298.50 | 0.0030 | 0.0003 | 0.0146 | 0.0017 | 94476 | 192 | 1377 | 468569 | 4232241 | **45** | 0.3102 |
| 30-34 | 81 | 22124.38 | 0.0037 | 0.0004 | 0.0181 | 0.0020 | 93099 | 246 | 1682 | 461367 | 3763672 | **40** | 0.3062 |
| 35-39 | 115 | 19804.26 | 0.0058 | 0.0005 | 0.0292 | 0.0027 | 91417 | 305 | 2666 | 451013 | 3302305 | **36** | 0.3018 |
| 40-44 | 129 | 16124.93 | 0.0080 | 0.0007 | 0.0395 | 0.0034 | 88751 | 384 | 3504 | 436007 | 2851292 | **32** | 0.2960 |
| 45-49 | 102 | 11724.15 | 0.0087 | 0.0009 | 0.0429 | 0.0042 | 85247 | 478 | 3661 | 417359 | 2415285 | **28** | 0.2885 |
| 50-54 | 112 | 10928.71 | 0.0102 | 0.0010 | 0.0496 | 0.0046 | 81586 | 579 | 4047 | 397882 | 1997926 | **24** | 0.2779 |
| 55-59 | 123 | 8905.13 | 0.0138 | 0.0013 | 0.0664 | 0.0058 | 77539 | 665 | 5152 | 375408 | 1600044 | **21** | 0.2704 |
| 60-64 | 243 | 8404.43 | 0.0289 | 0.0019 | 0.1381 | 0.0082 | 72387 | 766 | 9996 | 338426 | 1224636 | **17** | 0.2638 |
| 65-69 | 225 | 5577.00 | 0.0403 | 0.0027 | 0.1846 | 0.0111 | 62391 | 894 | 11518 | 282935 | 886210 | **14** | 0.2621 |
| 70-74 | 213 | 4079.31 | 0.0522 | 0.0036 | 0.2329 | 0.0140 | 50873 | 1008 | 11847 | 224820 | 603275 | **12** | 0.2548 |
| 75-79 | 160 | 2250.82 | 0.0711 | 0.0056 | 0.3007 | 0.0199 | 39026 | 1057 | 11737 | 165442 | 378455 | **10** | 0.2470 |
| 80-84 | 172 | 1818.09 | 0.0946 | 0.0072 | 0.3716 | 0.0225 | 27289 | 1072 | 10141 | 109261 | 213013 | **8** | 0.1961 |
| 85+ | 166 | 1199.33 | 0.1384 | 0.0108 | 1.0000 | 0.0000 | 17148 | 921 | 17148 | 103752 | 103752 | **6** | 0.0000 |

Table S4g. Lifetable for the period 2015-18 for females (conventional method)

| Age (years) | nDx | nPYx | nMx | SE nMx | nqx | SE nqx | lx | SE lx | ndx | nLx | Tx | ex (years) | SE ex (years) |
| --- | --- | --- | --- | --- | --- | --- | --- | --- | --- | --- | --- | --- | --- |
| <1 | 282 | 15872.04 | 0.0178 | 0.0011 | 0.0185 | 0.0011 | 100000 |  | 1853 | 98488 | 7359557 | **74** | 0.2800 |
| 1-4 | 112 | 67123.92 | 0.0017 | 0.0002 | 0.0067 | 0.0006 | 98147 | 109 | 654 | 391007 | 7261069 | **74** | 0.2729 |
| 5-9 | 75 | 87027.49 | 0.0009 | 0.0001 | 0.0043 | 0.0005 | 97493 | 124 | 421 | 486238 | 6870062 | **70** | 0.2708 |
| 10-14 | 39 | 78892.31 | 0.0005 | 0.0001 | 0.0025 | 0.0004 | 97072 | 133 | 240 | 484767 | 6383824 | **66** | 0.2698 |
| 15-19 | 45 | 56719.83 | 0.0008 | 0.0001 | 0.0039 | 0.0006 | 96832 | 138 | 382 | 483115 | 5899057 | **61** | 0.2693 |
| 20-24 | 59 | 43957.65 | 0.0013 | 0.0002 | 0.0066 | 0.0009 | 96450 | 149 | 640 | 480546 | 5415942 | **56** | 0.2682 |
| 25-29 | 89 | 37555.78 | 0.0024 | 0.0003 | 0.0118 | 0.0012 | 95810 | 170 | 1132 | 476364 | 4935396 | **52** | 0.2658 |
| 30-34 | 97 | 32887.24 | 0.0029 | 0.0003 | 0.0149 | 0.0015 | 94678 | 206 | 1409 | 469959 | 4459032 | **47** | 0.2617 |
| 35-39 | 115 | 29922.05 | 0.0038 | 0.0004 | 0.0189 | 0.0017 | 93269 | 248 | 1765 | 462230 | 3989073 | **43** | 0.2566 |
| 40-44 | 102 | 25362.59 | 0.0040 | 0.0004 | 0.0198 | 0.0019 | 91504 | 293 | 1813 | 452596 | 3526843 | **39** | 0.2512 |
| 45-49 | 111 | 21264.16 | 0.0052 | 0.0005 | 0.0256 | 0.0024 | 89691 | 338 | 2292 | 442534 | 3074247 | **34** | 0.2456 |
| 50-54 | 92 | 14749.99 | 0.0062 | 0.0007 | 0.0306 | 0.0031 | 87399 | 394 | 2673 | 430505 | 2631713 | **30** | 0.2389 |
| 55-59 | 136 | 18367.47 | 0.0074 | 0.0006 | 0.0360 | 0.0030 | 84726 | 471 | 3052 | 415955 | 2201208 | **26** | 0.2286 |
| 60-64 | 138 | 12111.68 | 0.0114 | 0.0010 | 0.0549 | 0.0045 | 81674 | 522 | 4481 | 396004 | 1785253 | **22** | 0.2244 |
| 65-69 | 189 | 9913.12 | 0.0191 | 0.0014 | 0.0928 | 0.0064 | 77193 | 619 | 7167 | 368524 | 1389249 | **18** | 0.2154 |
| 70-74 | 179 | 6742.74 | 0.0265 | 0.0020 | 0.1248 | 0.0087 | 70026 | 753 | 8742 | 326891 | 1020725 | **15** | 0.2046 |
| 75-79 | 224 | 4988.53 | 0.0449 | 0.0030 | 0.1951 | 0.0117 | 61284 | 904 | 11954 | 272232 | 693834 | **11** | 0.1876 |
| 80-84 | 155 | 2354.35 | 0.0658 | 0.0053 | 0.2849 | 0.0194 | 49330 | 1038 | 14055 | 211361 | 421602 | **9** | 0.1639 |
| 85+ | 237 | 2434.86 | 0.0973 | 0.0063 | 1.0000 | 0.0000 | 35275 | 1217 | 35275 | 210241 | 210241 | **6** | 0.0000 |

Table S4h. Lifetable for the period 2015-18 for males (conventional method)

| Age (years) | nDx | nPYx | nMx | SE nMx | nqx | SE nqx | lx | SE lx | ndx | nLx | Tx | ex (years) | SE ex (years) |
| --- | --- | --- | --- | --- | --- | --- | --- | --- | --- | --- | --- | --- | --- |
| <1 | 356 | 16323.15 | 0.0218 | 0.0012 | 0.0229 | 0.0012 | 100000 |  | 2285 | 98042 | 6782828 | **68** | 0.3028 |
| 1-4 | 161 | 69363.86 | 0.0023 | 0.0002 | 0.0093 | 0.0007 | 97715 | 119 | 908 | 388480 | 6684786 | **68** | 0.2981 |
| 5-9 | 89 | 88579.15 | 0.0010 | 0.0001 | 0.0050 | 0.0005 | 96807 | 138 | 485 | 482873 | 6296306 | **65** | 0.2967 |
| 10-14 | 72 | 80893.03 | 0.0009 | 0.0001 | 0.0044 | 0.0005 | 96322 | 146 | 425 | 480434 | 5813433 | **60** | 0.2963 |
| 15-19 | 72 | 59504.73 | 0.0012 | 0.0001 | 0.0062 | 0.0007 | 95897 | 154 | 595 | 478030 | 5332999 | **56** | 0.2961 |
| 20-24 | 53 | 38662.92 | 0.0014 | 0.0002 | 0.0069 | 0.0009 | 95302 | 169 | 656 | 474802 | 4854969 | **51** | 0.2954 |
| 25-29 | 70 | 29296.19 | 0.0024 | 0.0003 | 0.0120 | 0.0014 | 94646 | 190 | 1137 | 470718 | 4380167 | **46** | 0.2938 |
| 30-34 | 79 | 24145.76 | 0.0033 | 0.0004 | 0.0163 | 0.0018 | 93509 | 231 | 1527 | 463698 | 3909449 | **42** | 0.2905 |
| 35-39 | 94 | 21666.48 | 0.0043 | 0.0005 | 0.0215 | 0.0022 | 91982 | 285 | 1980 | 455259 | 3445751 | **37** | 0.2859 |
| 40-44 | 106 | 18321.62 | 0.0058 | 0.0006 | 0.0286 | 0.0027 | 90002 | 344 | 2574 | 443466 | 2990492 | **33** | 0.2810 |
| 45-49 | 102 | 14604.63 | 0.0070 | 0.0007 | 0.0347 | 0.0034 | 87428 | 416 | 3034 | 429583 | 2547026 | **29** | 0.2752 |
| 50-54 | 117 | 11128.57 | 0.0105 | 0.0010 | 0.0512 | 0.0046 | 84394 | 499 | 4324 | 411993 | 2117443 | **25** | 0.2682 |
| 55-59 | 136 | 10091.33 | 0.0135 | 0.0012 | 0.0655 | 0.0054 | 80070 | 613 | 5244 | 387430 | 1705450 | **21** | 0.2584 |
| 60-64 | 205 | 8771.08 | 0.0234 | 0.0016 | 0.1109 | 0.0073 | 74826 | 721 | 8297 | 355585 | 1318020 | **18** | 0.2507 |
| 65-69 | 252 | 6804.71 | 0.0370 | 0.0023 | 0.1691 | 0.0097 | 66529 | 842 | 11247 | 302453 | 962435 | **14** | 0.2463 |
| 70-74 | 215 | 4448.51 | 0.0483 | 0.0033 | 0.2160 | 0.0130 | 55282 | 959 | 11941 | 244702 | 659982 | **12** | 0.2423 |
| 75-79 | 199 | 2923.56 | 0.0681 | 0.0048 | 0.2969 | 0.0176 | 43341 | 1047 | 12867 | 182803 | 415280 | **10** | 0.2324 |
| 80-84 | 149 | 1539.07 | 0.0968 | 0.0080 | 0.3905 | 0.0250 | 30474 | 1080 | 11900 | 123473 | 232477 | **8** | 0.2072 |
| 85+ | 236 | 1610.53 | 0.1465 | 0.0096 | 1.0000 | 0.0000 | 18574 | 1006 | 18574 | 109004 | 109004 | **6** | 0.0000 |

Table S5a. Lifetable for the period 2003-06 for females (INDEPTH - MADIMAH method)

| Age (years) | nDx | nPYx | nMx | SE nMx | nqx | SE nqx | lx | SE lx | ndx | nLx | Tx | ex (years) | SE ex (years) |
| --- | --- | --- | --- | --- | --- | --- | --- | --- | --- | --- | --- | --- | --- |
| <1 | 602 | 17879.49 | 0.0337 | 0.0014 | 0.0334 | 0.0013 | 100000 |  | 3343 | 97441 | 6484412 | **65** | 0.3277 |
| 1-4 | 369 | 67540.14 | 0.0055 | 0.0003 | 0.0213 | 0.0011 | 96657 | 133 | 2056 | 381212 | 6386971 | **66** | 0.3261 |
| 5-9 | 101 | 73700.32 | 0.0014 | 0.0001 | 0.0067 | 0.0007 | 94601 | 168 | 636 | 471113 | 6005759 | **63** | 0.3249 |
| 10-14 | 53 | 62049.61 | 0.0009 | 0.0001 | 0.0044 | 0.0006 | 93965 | 178 | 412 | 468800 | 5534646 | **59** | 0.3245 |
| 15-19 | 73 | 50447.76 | 0.0014 | 0.0002 | 0.0072 | 0.0008 | 93553 | 186 | 678 | 466209 | 5065846 | **54** | 0.3241 |
| 20-24 | 124 | 37020.91 | 0.0033 | 0.0003 | 0.0169 | 0.0015 | 92875 | 201 | 1567 | 460577 | 4599637 | **50** | 0.3235 |
| 25-29 | 201 | 35310.63 | 0.0057 | 0.0004 | 0.0283 | 0.0020 | 91308 | 242 | 2587 | 449846 | 4139060 | **45** | 0.3208 |
| 30-34 | 202 | 26823.56 | 0.0075 | 0.0005 | 0.0370 | 0.0026 | 88721 | 296 | 3283 | 435408 | 3689214 | **42** | 0.3178 |
| 35-39 | 176 | 20911.50 | 0.0084 | 0.0006 | 0.0413 | 0.0030 | 85438 | 364 | 3528 | 417113 | 3253806 | **38** | 0.3119 |
| 40-44 | 139 | 18511.24 | 0.0075 | 0.0006 | 0.0372 | 0.0031 | 81910 | 439 | 3049 | 401903 | 2836693 | **35** | 0.3028 |
| 45-49 | 145 | 17942.51 | 0.0081 | 0.0007 | 0.0391 | 0.0032 | 78861 | 494 | 3080 | 385589 | 2434790 | **31** | 0.2956 |
| 50-54 | 104 | 12458.03 | 0.0083 | 0.0008 | 0.0416 | 0.0040 | 75781 | 539 | 3152 | 371042 | 2049201 | **27** | 0.2913 |
| 55-59 | 127 | 10695.48 | 0.0119 | 0.0011 | 0.0556 | 0.0048 | 72629 | 598 | 4037 | 351871 | 1678159 | **23** | 0.2846 |
| 60-64 | 162 | 8676.49 | 0.0187 | 0.0015 | 0.0901 | 0.0067 | 68592 | 667 | 6177 | 328858 | 1326288 | **19** | 0.2796 |
| 65-69 | 161 | 5888.15 | 0.0273 | 0.0022 | 0.1328 | 0.0097 | 62415 | 763 | 8286 | 288263 | 997430 | **16** | 0.2758 |
| 70-74 | 158 | 4256.17 | 0.0371 | 0.0030 | 0.1678 | 0.0122 | 54129 | 910 | 9085 | 247300 | 709167 | **13** | 0.2626 |
| 75-79 | 132 | 2390.65 | 0.0552 | 0.0048 | 0.2478 | 0.0187 | 45044 | 1009 | 11162 | 191272 | 461867 | **10** | 0.2536 |
| 80-84 | 86 | 1295.86 | 0.0664 | 0.0072 | 0.2852 | 0.0260 | 33882 | 1197 | 9663 | 146163 | 270595 | **8** | 0.1961 |
| 85+ | 89 | 837.56 | 0.1063 | 0.0113 | 1.0000 | 0.0000 | 24219 | 1230 | 24219 | 124432 | 124432 | **5** | 0.0000 |

Table S5b. Lifetable for the period 2003-06 for males (INDEPTH - MADIMAH method)

| Age (years) | nDx | nPYx | nMx | SE nMx | nqx | SE nqx | lx | SE lx | ndx | nLx | Tx | ex (years) | SE ex (years) |
| --- | --- | --- | --- | --- | --- | --- | --- | --- | --- | --- | --- | --- | --- |
| <1 | 712 | 18169.75 | 0.0392 | 0.0015 | 0.0389 | 0.0014 | 100000 |  | 3889 | 96851 | 6129690 | **61** | 0.3436 |
| 1-4 | 393 | 68556.24 | 0.0057 | 0.0003 | 0.0224 | 0.0011 | 96111 | 142 | 2156 | 379103 | 6032839 | **63** | 0.3448 |
| 5-9 | 141 | 74170.68 | 0.0019 | 0.0002 | 0.0093 | 0.0008 | 93955 | 176 | 875 | 467189 | 5653736 | **60** | 0.3453 |
| 10-14 | 75 | 64415.36 | 0.0012 | 0.0001 | 0.0059 | 0.0007 | 93080 | 189 | 546 | 463887 | 5186547 | **56** | 0.3455 |
| 15-19 | 82 | 50648.46 | 0.0016 | 0.0002 | 0.0082 | 0.0009 | 92534 | 198 | 757 | 460749 | 4722660 | **51** | 0.3456 |
| 20-24 | 61 | 28961.99 | 0.0021 | 0.0003 | 0.0107 | 0.0014 | 91777 | 214 | 983 | 456415 | 4261911 | **46** | 0.3456 |
| 25-29 | 95 | 23775.69 | 0.0040 | 0.0004 | 0.0199 | 0.0020 | 90794 | 246 | 1811 | 449039 | 3805496 | **42** | 0.3439 |
| 30-34 | 124 | 19396.97 | 0.0064 | 0.0006 | 0.0320 | 0.0028 | 88983 | 303 | 2850 | 437416 | 3356457 | **38** | 0.3408 |
| 35-39 | 143 | 14909.60 | 0.0096 | 0.0008 | 0.0462 | 0.0038 | 86133 | 385 | 3980 | 419658 | 2919041 | **34** | 0.3355 |
| 40-44 | 113 | 12748.02 | 0.0089 | 0.0008 | 0.0436 | 0.0040 | 82153 | 495 | 3583 | 401659 | 2499383 | **30** | 0.3264 |
| 45-49 | 145 | 10457.39 | 0.0140 | 0.0012 | 0.0659 | 0.0053 | 78570 | 576 | 5177 | 378698 | 2097724 | **27** | 0.3184 |
| 50-54 | 162 | 9632.97 | 0.0169 | 0.0013 | 0.0840 | 0.0063 | 73393 | 682 | 6165 | 353121 | 1719026 | **23** | 0.3073 |
| 55-59 | 134 | 7687.03 | 0.0176 | 0.0015 | 0.0822 | 0.0068 | 67228 | 773 | 5526 | 321983 | 1365905 | **20** | 0.2975 |
| 60-64 | 170 | 6222.93 | 0.0273 | 0.0021 | 0.1295 | 0.0093 | 61702 | 848 | 7991 | 290785 | 1043922 | **17** | 0.2905 |
| 65-69 | 173 | 4303.51 | 0.0404 | 0.0031 | 0.1866 | 0.0128 | 53711 | 933 | 10024 | 242169 | 753137 | **14** | 0.2854 |
| 70-74 | 169 | 3404.14 | 0.0494 | 0.0038 | 0.2199 | 0.0150 | 43687 | 1028 | 9606 | 195579 | 510968 | **12** | 0.2694 |
| 75-79 | 151 | 2058.37 | 0.0734 | 0.0060 | 0.3044 | 0.0207 | 34081 | 1033 | 10373 | 139104 | 315389 | **9** | 0.2633 |
| 80-84 | 105 | 1199.73 | 0.0875 | 0.0086 | 0.3583 | 0.0280 | 23708 | 1041 | 8495 | 97079 | 176285 | **7** | 0.2166 |
| 85+ | 84 | 836.45 | 0.1004 | 0.0110 | 1.0000 | 0.0000 | 15213 | 935 | 15213 | 79206 | 79206 | **5** | 0.0000 |

Table S5c. Lifetable for the period 2007-10 for females (INDEPTH - MADIMAH method)

| Age (years) | nDx | nPYx | nMx | SE nMx | nqx | SE nqx | lx | SE lx | ndx | nLx | Tx | ex (years) | SE ex (years) |
| --- | --- | --- | --- | --- | --- | --- | --- | --- | --- | --- | --- | --- | --- |
| <1 | 384 | 18892.12 | 0.0203 | 0.0010 | 0.0202 | 0.0010 | 100000 |  | 2020 | 98419 | 7016073 | **70** | 0.2977 |
| 1-4 | 183 | 74887.49 | 0.0024 | 0.0002 | 0.0096 | 0.0007 | 97980 | 102 | 940 | 389547 | 6917654 | **71** | 0.2945 |
| 5-9 | 65 | 83665.15 | 0.0008 | 0.0001 | 0.0039 | 0.0005 | 97040 | 122 | 375 | 484237 | 6528107 | **67** | 0.2932 |
| 10-14 | 45 | 72645.63 | 0.0006 | 0.0001 | 0.0031 | 0.0005 | 96665 | 130 | 301 | 482585 | 6043870 | **63** | 0.2927 |
| 15-19 | 55 | 56809.09 | 0.0010 | 0.0001 | 0.0048 | 0.0007 | 96364 | 137 | 467 | 480625 | 5561285 | **58** | 0.2923 |
| 20-24 | 81 | 46111.60 | 0.0018 | 0.0002 | 0.0089 | 0.0010 | 95897 | 150 | 856 | 477516 | 5080660 | **53** | 0.2914 |
| 25-29 | 108 | 34989.33 | 0.0031 | 0.0003 | 0.0155 | 0.0015 | 95041 | 177 | 1471 | 472031 | 4603144 | **48** | 0.2897 |
| 30-34 | 140 | 33148.43 | 0.0042 | 0.0004 | 0.0210 | 0.0018 | 93570 | 223 | 1969 | 462866 | 4131113 | **44** | 0.2859 |
| 35-39 | 137 | 25514.92 | 0.0054 | 0.0005 | 0.0267 | 0.0022 | 91601 | 274 | 2445 | 452281 | 3668247 | **40** | 0.2818 |
| 40-44 | 94 | 18340.43 | 0.0051 | 0.0005 | 0.0263 | 0.0027 | 89156 | 337 | 2345 | 439311 | 3215966 | **36** | 0.2756 |
| 45-49 | 103 | 19494.60 | 0.0053 | 0.0005 | 0.0263 | 0.0026 | 86811 | 406 | 2287 | 428056 | 2776655 | **32** | 0.2665 |
| 50-54 | 105 | 16205.93 | 0.0065 | 0.0006 | 0.0318 | 0.0031 | 84524 | 455 | 2692 | 415417 | 2348599 | **28** | 0.2617 |
| 55-59 | 145 | 12256.09 | 0.0118 | 0.0010 | 0.0583 | 0.0047 | 81832 | 513 | 4772 | 398691 | 1933182 | **24** | 0.2572 |
| 60-64 | 148 | 9917.64 | 0.0149 | 0.0012 | 0.0751 | 0.0059 | 77060 | 615 | 5786 | 369828 | 1534491 | **20** | 0.2499 |
| 65-69 | 204 | 8080.71 | 0.0252 | 0.0018 | 0.1167 | 0.0077 | 71274 | 726 | 8318 | 334491 | 1164663 | **16** | 0.2411 |
| 70-74 | 151 | 4514.11 | 0.0335 | 0.0027 | 0.1561 | 0.0117 | 62956 | 849 | 9825 | 286954 | 830172 | **13** | 0.2358 |
| 75-79 | 188 | 3603.00 | 0.0522 | 0.0038 | 0.2322 | 0.0148 | 53131 | 1038 | 12337 | 235589 | 543218 | **10** | 0.2132 |
| 80-84 | 100 | 1400.36 | 0.0714 | 0.0072 | 0.3052 | 0.0254 | 40794 | 1120 | 12452 | 167673 | 307629 | **8** | 0.1998 |
| 85+ | 138 | 1323.77 | 0.1042 | 0.0089 | 1.0000 | 0.0000 | 28342 | 1370 | 28342 | 139956 | 139956 | **5** | 0.0000 |

Table S5d. Lifetable for the period 2007-10 for males (INDEPTH - MADIMAH method)

| Age (years) | nDx | nPYx | nMx | SE nMx | nqx | SE nqx | lx | SE lx | ndx | nLx | Tx | ex (years) | SE ex (years) |
| --- | --- | --- | --- | --- | --- | --- | --- | --- | --- | --- | --- | --- | --- |
| <1 | 470 | 19203.04 | 0.0244 | 0.0011 | 0.0243 | 0.0011 | 100000 |  | 2432 | 98034 | 6640873 | **66** | 0.3203 |
| 1-4 | 224 | 75380.74 | 0.0030 | 0.0002 | 0.0117 | 0.0008 | 97568 | 110 | 1137 | 387434 | 6542839 | **67** | 0.3191 |
| 5-9 | 79 | 84799.19 | 0.0009 | 0.0001 | 0.0046 | 0.0005 | 96431 | 133 | 447 | 480976 | 6155405 | **64** | 0.3187 |
| 10-14 | 59 | 73928.62 | 0.0008 | 0.0001 | 0.0039 | 0.0005 | 95984 | 141 | 379 | 478853 | 5674429 | **59** | 0.3185 |
| 15-19 | 64 | 57842.92 | 0.0011 | 0.0001 | 0.0056 | 0.0007 | 95605 | 149 | 534 | 476709 | 5195576 | **54** | 0.3185 |
| 20-24 | 58 | 36392.13 | 0.0016 | 0.0002 | 0.0081 | 0.0011 | 95071 | 163 | 768 | 473375 | 4718867 | **50** | 0.3182 |
| 25-29 | 67 | 25561.14 | 0.0026 | 0.0003 | 0.0132 | 0.0016 | 94303 | 191 | 1248 | 468778 | 4245492 | **45** | 0.3167 |
| 30-34 | 95 | 23246.27 | 0.0041 | 0.0004 | 0.0204 | 0.0021 | 93055 | 240 | 1902 | 460335 | 3776714 | **41** | 0.3133 |
| 35-39 | 108 | 18860.83 | 0.0057 | 0.0006 | 0.0281 | 0.0027 | 91153 | 305 | 2565 | 449545 | 3316379 | **36** | 0.3090 |
| 40-44 | 88 | 14137.36 | 0.0062 | 0.0007 | 0.0309 | 0.0032 | 88588 | 384 | 2736 | 435626 | 2866834 | **32** | 0.3033 |
| 45-49 | 98 | 12563.73 | 0.0077 | 0.0008 | 0.0393 | 0.0039 | 85852 | 471 | 3371 | 421056 | 2431208 | **28** | 0.2953 |
| 50-54 | 116 | 9982.17 | 0.0115 | 0.0011 | 0.0549 | 0.0050 | 82481 | 560 | 4527 | 400843 | 2010152 | **24** | 0.2873 |
| 55-59 | 161 | 9662.16 | 0.0166 | 0.0013 | 0.0791 | 0.0060 | 77954 | 674 | 6163 | 374522 | 1609309 | **21** | 0.2784 |
| 60-64 | 166 | 6958.68 | 0.0239 | 0.0019 | 0.1147 | 0.0084 | 71791 | 777 | 8235 | 338562 | 1234787 | **17** | 0.2738 |
| 65-69 | 187 | 5404.20 | 0.0344 | 0.0025 | 0.1599 | 0.0107 | 63556 | 912 | 10165 | 293305 | 896225 | **14** | 0.2666 |
| 70-74 | 180 | 3282.33 | 0.0548 | 0.0041 | 0.2453 | 0.0159 | 53391 | 1026 | 13097 | 230927 | 602920 | **11** | 0.2645 |
| 75-79 | 200 | 2749.55 | 0.0727 | 0.0052 | 0.3000 | 0.0177 | 40294 | 1150 | 12088 | 169955 | 371993 | **9** | 0.2434 |
| 80-84 | 115 | 1204.00 | 0.0955 | 0.0090 | 0.3841 | 0.0281 | 28206 | 1081 | 10834 | 106528 | 202038 | **7** | 0.2441 |
| 85+ | 140 | 1172.58 | 0.1194 | 0.0101 | 1.0000 | 0.0000 | 17372 | 1054 | 17372 | 95510 | 95510 | **5** | 0.0000 |

Table S5e. Lifetable for the period 2011-14 for females (INDEPTH - MADIMAH method)

| Age (years) | nDx | nPYx | nMx | SE nMx | nqx | SE nqx | lx | SE lx | ndx | nLx | Tx | ex (years) | SE ex (years) |
| --- | --- | --- | --- | --- | --- | --- | --- | --- | --- | --- | --- | --- | --- |
| <1 | 355 | 18535.20 | 0.0191 | 0.0010 | 0.0193 | 0.0010 | 100000 |  | 1929 | 98416 | 7264656 | **73** | 0.2920 |
| 1-4 | 174 | 75544.98 | 0.0023 | 0.0002 | 0.0092 | 0.0007 | 98071 | 101 | 900 | 390052 | 7166240 | **73** | 0.2878 |
| 5-9 | 80 | 88990.54 | 0.0009 | 0.0001 | 0.0044 | 0.0005 | 97171 | 121 | 432 | 484606 | 6776188 | **70** | 0.2860 |
| 10-14 | 42 | 79433.92 | 0.0005 | 0.0001 | 0.0026 | 0.0004 | 96739 | 130 | 254 | 482999 | 6291582 | **65** | 0.2853 |
| 15-19 | 57 | 61744.36 | 0.0009 | 0.0001 | 0.0046 | 0.0006 | 96485 | 135 | 444 | 481254 | 5808583 | **60** | 0.2849 |
| 20-24 | 63 | 48336.67 | 0.0013 | 0.0002 | 0.0066 | 0.0008 | 96041 | 147 | 632 | 478812 | 5327329 | **55** | 0.2840 |
| 25-29 | 80 | 40438.39 | 0.0020 | 0.0002 | 0.0099 | 0.0011 | 95409 | 166 | 945 | 474642 | 4848517 | **51** | 0.2825 |
| 30-34 | 116 | 33011.72 | 0.0035 | 0.0003 | 0.0172 | 0.0016 | 94464 | 195 | 1624 | 468587 | 4373875 | **46** | 0.2801 |
| 35-39 | 113 | 29688.01 | 0.0038 | 0.0004 | 0.0192 | 0.0018 | 92840 | 243 | 1781 | 459531 | 3905288 | **42** | 0.2759 |
| 40-44 | 100 | 23891.64 | 0.0042 | 0.0004 | 0.0210 | 0.0021 | 91059 | 291 | 1912 | 450144 | 3445757 | **38** | 0.2714 |
| 45-49 | 79 | 16147.82 | 0.0049 | 0.0006 | 0.0255 | 0.0028 | 89147 | 342 | 2273 | 440332 | 2995613 | **34** | 0.2663 |
| 50-54 | 135 | 19736.80 | 0.0068 | 0.0006 | 0.0327 | 0.0028 | 86874 | 416 | 2842 | 427038 | 2555281 | **29** | 0.2572 |
| 55-59 | 135 | 14182.96 | 0.0095 | 0.0008 | 0.0471 | 0.0040 | 84032 | 471 | 3958 | 409272 | 2128243 | **25** | 0.2536 |
| 60-64 | 164 | 11434.12 | 0.0143 | 0.0011 | 0.0690 | 0.0052 | 80074 | 560 | 5523 | 387079 | 1718971 | **21** | 0.2464 |
| 65-69 | 163 | 8379.15 | 0.0195 | 0.0015 | 0.0959 | 0.0071 | 74551 | 669 | 7150 | 354856 | 1331892 | **18** | 0.2393 |
| 70-74 | 230 | 6760.38 | 0.0340 | 0.0022 | 0.1583 | 0.0096 | 67401 | 808 | 10667 | 309916 | 977036 | **14** | 0.2280 |
| 75-79 | 145 | 3196.06 | 0.0454 | 0.0038 | 0.2032 | 0.0151 | 56734 | 948 | 11530 | 252186 | 667120 | **12** | 0.2167 |
| 80-84 | 137 | 2711.97 | 0.0505 | 0.0043 | 0.2302 | 0.0173 | 45204 | 1151 | 10407 | 198756 | 414934 | **9** | 0.1524 |
| 85+ | 132 | 1503.15 | 0.0878 | 0.0077 | 1.0000 | 0.0000 | 34797 | 1210 | 34797 | 216178 | 216178 | **6** | 0.0000 |

Table S5f. Lifetable for the period 2011-14 for males (INDEPTH - MADIMAH method)

| Age (years) | nDx | nPYx | nMx | SE nMx | nqx | SE nqx | lx | SE lx | ndx | nLx | Tx | ex (years) | SE ex (years) |
| --- | --- | --- | --- | --- | --- | --- | --- | --- | --- | --- | --- | --- | --- |
| <1 | 405 | 19070.32 | 0.0212 | 0.0011 | 0.0213 | 0.0010 | 100000 |  | 2131 | 98263 | 6707473 | **67** | 0.3070 |
| 1-4 | 198 | 77034.44 | 0.0026 | 0.0002 | 0.0103 | 0.0007 | 97869 | 104 | 1006 | 388982 | 6609210 | **68** | 0.3051 |
| 5-9 | 97 | 90443.23 | 0.0011 | 0.0001 | 0.0053 | 0.0005 | 96863 | 125 | 512 | 482966 | 6220228 | **64** | 0.3043 |
| 10-14 | 62 | 81074.22 | 0.0008 | 0.0001 | 0.0038 | 0.0005 | 96351 | 135 | 367 | 480787 | 5737262 | **60** | 0.3041 |
| 15-19 | 50 | 62108.66 | 0.0008 | 0.0001 | 0.0042 | 0.0006 | 95984 | 142 | 400 | 479031 | 5256475 | **55** | 0.3040 |
| 20-24 | 55 | 40526.01 | 0.0014 | 0.0002 | 0.0069 | 0.0009 | 95584 | 153 | 655 | 476258 | 4777444 | **50** | 0.3037 |
| 25-29 | 75 | 29202.84 | 0.0026 | 0.0003 | 0.0127 | 0.0015 | 94929 | 175 | 1201 | 471277 | 4301186 | **45** | 0.3026 |
| 30-34 | 81 | 24854.63 | 0.0033 | 0.0004 | 0.0163 | 0.0018 | 93728 | 222 | 1527 | 464826 | 3829909 | **41** | 0.2996 |
| 35-39 | 115 | 21657.86 | 0.0053 | 0.0005 | 0.0268 | 0.0025 | 92201 | 275 | 2472 | 455249 | 3365083 | **36** | 0.2962 |
| 40-44 | 129 | 17467.97 | 0.0074 | 0.0007 | 0.0373 | 0.0032 | 89729 | 351 | 3349 | 441048 | 2909834 | **32** | 0.2913 |
| 45-49 | 102 | 12506.66 | 0.0082 | 0.0008 | 0.0397 | 0.0038 | 86380 | 444 | 3425 | 423378 | 2468786 | **29** | 0.2844 |
| 50-54 | 112 | 11555.13 | 0.0097 | 0.0009 | 0.0471 | 0.0043 | 82955 | 543 | 3905 | 404804 | 2045408 | **25** | 0.2755 |
| 55-59 | 123 | 9354.78 | 0.0131 | 0.0012 | 0.0633 | 0.0055 | 79050 | 631 | 5005 | 382927 | 1640604 | **21** | 0.2689 |
| 60-64 | 243 | 8735.48 | 0.0278 | 0.0018 | 0.1357 | 0.0081 | 74045 | 735 | 10051 | 345742 | 1257677 | **17** | 0.2631 |
| 65-69 | 225 | 5760.74 | 0.0391 | 0.0026 | 0.1814 | 0.0109 | 63994 | 876 | 11607 | 289675 | 911935 | **14** | 0.2613 |
| 70-74 | 213 | 4199.40 | 0.0507 | 0.0035 | 0.2342 | 0.0140 | 52387 | 1005 | 12267 | 230149 | 622260 | **12** | 0.2537 |
| 75-79 | 160 | 2305.71 | 0.0694 | 0.0055 | 0.2857 | 0.0191 | 40120 | 1070 | 11461 | 170419 | 392111 | **10** | 0.2415 |
| 80-84 | 172 | 1863.27 | 0.0923 | 0.0071 | 0.3662 | 0.0222 | 28659 | 1095 | 10494 | 114006 | 221692 | **8** | 0.1938 |
| 85+ | 166 | 1225.15 | 0.1355 | 0.0106 | 1.0000 | 0.0000 | 18165 | 954 | 18165 | 107686 | 107686 | **6** | 0.0000 |

Table S5g. Lifetable for the period 2015-18 for females (INDEPTH - MADIMAH method)

| Age (years) | nDx | nPYx | nMx | SE nMx | nqx | SE nqx | lx | SE lx | ndx | nLx | Tx | ex (years) | SE ex (years) |
| --- | --- | --- | --- | --- | --- | --- | --- | --- | --- | --- | --- | --- | --- |
| <1 | 282 | 17495.56 | 0.0161 | 0.0010 | 0.0168 | 0.0010 | 100000 |  | 1682 | 98627 | 7368025 | **74** | 0.2765 |
| 1-4 | 112 | 71776.81 | 0.0016 | 0.0001 | 0.0066 | 0.0006 | 98318 | 97 | 650 | 391698 | 7269398 | **74** | 0.2709 |
| 5-9 | 75 | 91731.29 | 0.0008 | 0.0001 | 0.0041 | 0.0005 | 97668 | 113 | 399 | 487174 | 6877700 | **70** | 0.2689 |
| 10-14 | 39 | 82867.75 | 0.0005 | 0.0001 | 0.0025 | 0.0004 | 97269 | 122 | 241 | 485746 | 6390526 | **66** | 0.2680 |
| 15-19 | 45 | 61877.45 | 0.0007 | 0.0001 | 0.0040 | 0.0006 | 97028 | 127 | 384 | 484081 | 5904780 | **61** | 0.2675 |
| 20-24 | 59 | 49695.10 | 0.0012 | 0.0002 | 0.0061 | 0.0008 | 96644 | 138 | 589 | 481634 | 5420699 | **56** | 0.2663 |
| 25-29 | 89 | 41948.68 | 0.0021 | 0.0002 | 0.0111 | 0.0012 | 96055 | 157 | 1062 | 477716 | 4939065 | **51** | 0.2644 |
| 30-34 | 97 | 35925.92 | 0.0027 | 0.0003 | 0.0140 | 0.0014 | 94993 | 190 | 1329 | 471670 | 4461349 | **47** | 0.2610 |
| 35-39 | 115 | 31887.99 | 0.0036 | 0.0003 | 0.0187 | 0.0017 | 93664 | 230 | 1753 | 464144 | 3989679 | **43** | 0.2567 |
| 40-44 | 102 | 26613.84 | 0.0038 | 0.0004 | 0.0196 | 0.0019 | 91911 | 275 | 1797 | 454573 | 3525535 | **38** | 0.2516 |
| 45-49 | 111 | 22193.00 | 0.0050 | 0.0005 | 0.0247 | 0.0023 | 90114 | 322 | 2230 | 444667 | 3070962 | **34** | 0.2463 |
| 50-54 | 92 | 15257.82 | 0.0060 | 0.0006 | 0.0316 | 0.0032 | 87884 | 377 | 2778 | 432479 | 2626295 | **30** | 0.2404 |
| 55-59 | 136 | 18899.11 | 0.0072 | 0.0006 | 0.0357 | 0.0030 | 85106 | 458 | 3037 | 417659 | 2193816 | **26** | 0.2294 |
| 60-64 | 138 | 12469.59 | 0.0111 | 0.0009 | 0.0540 | 0.0045 | 82069 | 510 | 4430 | 397750 | 1776157 | **22** | 0.2256 |
| 65-69 | 189 | 10193.08 | 0.0185 | 0.0014 | 0.0959 | 0.0066 | 77639 | 607 | 7446 | 369400 | 1378407 | **18** | 0.2178 |
| 70-74 | 179 | 6983.94 | 0.0256 | 0.0019 | 0.1294 | 0.0090 | 70193 | 745 | 9084 | 325935 | 1009007 | **14** | 0.2065 |
| 75-79 | 224 | 5180.00 | 0.0432 | 0.0029 | 0.1975 | 0.0118 | 61109 | 897 | 12072 | 269770 | 683072 | **11** | 0.1881 |
| 80-84 | 155 | 2441.06 | 0.0635 | 0.0051 | 0.2848 | 0.0193 | 49037 | 1034 | 13968 | 208569 | 413302 | **8** | 0.1637 |
| 85+ | 237 | 2560.76 | 0.0926 | 0.0060 | 1.0000 | 0.0000 | 35069 | 1198 | 35069 | 204733 | 204733 | **6** | 0.0000 |

Table S5h. Lifetable for the period 2015-18 for males (INDEPTH - MADIMAH method)

| Age (years) | nDx | nPYx | nMx | SE nMx | nqx | SE nqx | lx | SE lx | ndx | nLx | Tx | ex (years) | SE ex (years) |
| --- | --- | --- | --- | --- | --- | --- | --- | --- | --- | --- | --- | --- | --- |
| <1 | 356 | 18049.69 | 0.0197 | 0.0010 | 0.0202 | 0.0011 | 100000 |  | 2023 | 98266 | 6824145 | **68** | 0.2966 |
| 1-4 | 161 | 74054.46 | 0.0022 | 0.0002 | 0.0089 | 0.0007 | 97977 | 105 | 871 | 389623 | 6725879 | **69** | 0.2932 |
| 5-9 | 89 | 93163.20 | 0.0010 | 0.0001 | 0.0049 | 0.0005 | 97106 | 124 | 473 | 484392 | 6336256 | **65** | 0.2919 |
| 10-14 | 72 | 84648.04 | 0.0009 | 0.0001 | 0.0044 | 0.0005 | 96633 | 133 | 430 | 481968 | 5851864 | **61** | 0.2915 |
| 15-19 | 72 | 63489.91 | 0.0011 | 0.0001 | 0.0058 | 0.0007 | 96203 | 141 | 561 | 479611 | 5369896 | **56** | 0.2912 |
| 20-24 | 53 | 43076.23 | 0.0012 | 0.0002 | 0.0068 | 0.0009 | 95642 | 155 | 654 | 476488 | 4890285 | **51** | 0.2906 |
| 25-29 | 70 | 33235.11 | 0.0021 | 0.0003 | 0.0109 | 0.0013 | 94988 | 176 | 1035 | 472615 | 4413797 | **46** | 0.2889 |
| 30-34 | 79 | 27084.94 | 0.0029 | 0.0003 | 0.0155 | 0.0017 | 93953 | 212 | 1454 | 466038 | 3941182 | **42** | 0.2863 |
| 35-39 | 94 | 23698.42 | 0.0040 | 0.0004 | 0.0201 | 0.0021 | 92499 | 262 | 1861 | 458030 | 3475144 | **38** | 0.2821 |
| 40-44 | 106 | 19780.58 | 0.0054 | 0.0005 | 0.0265 | 0.0025 | 90638 | 318 | 2406 | 446936 | 3017114 | **33** | 0.2780 |
| 45-49 | 102 | 15718.03 | 0.0065 | 0.0006 | 0.0331 | 0.0032 | 88232 | 387 | 2924 | 433691 | 2570178 | **29** | 0.2733 |
| 50-54 | 117 | 11845.05 | 0.0099 | 0.0009 | 0.0504 | 0.0045 | 85308 | 468 | 4302 | 416310 | 2136487 | **25** | 0.2672 |
| 55-59 | 136 | 10601.13 | 0.0128 | 0.0011 | 0.0653 | 0.0054 | 81006 | 584 | 5292 | 391583 | 1720177 | **21** | 0.2577 |
| 60-64 | 205 | 9126.99 | 0.0225 | 0.0016 | 0.1125 | 0.0074 | 75714 | 697 | 8520 | 358809 | 1328594 | **18** | 0.2500 |
| 65-69 | 252 | 7049.09 | 0.0357 | 0.0023 | 0.1637 | 0.0094 | 67194 | 825 | 11001 | 305445 | 969785 | **14** | 0.2446 |
| 70-74 | 215 | 4582.65 | 0.0469 | 0.0032 | 0.2162 | 0.0131 | 56193 | 944 | 12148 | 247528 | 664340 | **12** | 0.2417 |
| 75-79 | 199 | 3007.35 | 0.0662 | 0.0047 | 0.2945 | 0.0175 | 44045 | 1039 | 12971 | 184702 | 416812 | **9** | 0.2318 |
| 80-84 | 149 | 1576.27 | 0.0945 | 0.0078 | 0.3955 | 0.0252 | 31074 | 1079 | 12290 | 124164 | 232110 | **7** | 0.2087 |
| 85+ | 236 | 1653.80 | 0.1427 | 0.0093 | 1.0000 | 0.0000 | 18784 | 1006 | 18784 | 107946 | 107946 | **6** | 0.0000 |

Table S6. Seasonality – high and low mortality months for each age group

| **Age group** | **Peak** | **Trough** |
| --- | --- | --- |
| 0-28 days | February | November |
| 29-365 days | June | January |
| 1-4 years | July | February |
| 5-14 years | January | February |
| 15-54 years | June | January |
| 55-74 years | June | January |
| 75+ years | August | January |

Table S7. Median age at death in years (based on the Kaplan-Meier method) for each location by period.

| **Location** | **2003-06** | | **2007-10** | | **2011-14** | | **2015-18** | | **^*^2003-2018** | |
| --- | --- | --- | --- | --- | --- | --- | --- | --- | --- | --- |
|  | female | male | female | male | female | male | female | male | **female**  **SD** | **male**  **SD** |
| Banda ra salam | 73 | 68 | 75 | 70 | 77 | 74 | 82 | 71 | 4 | 3 |
| Chasimba | 75 | 66 | 78 | 69 | 85 | 72 | 80 | 70 | 4 | 3 |
| Gede | 73 | 70 | 79 | 69 | 79 | 72 | 77 | 71 | 3 | 1 |
| Jaribuni | 77 | 68 | 78 | 71 | 74 | 69 | 80 | 75 | 3 | 3 |
| Junju | 67 | 64 | 73 | 69 | 76 | 69 | 78 | 71 | 5 | 3 |
| Kauma | 80 | 66 | 74 | 70 | 79 | 68 | 82 | 72 | 3 | 3 |
| Kilifi township | 70 | 67 | 74 | 73 | 77 | 73 | 78 | 73 | 4 | 3 |
| Matsangoni | 72 | 71 | 78 | 68 | 78 | 70 | 78 | 73 | 3 | 2 |
| Mtwapa | 70 | 70 | 75 | 74 | 75 | 70 | 81 | 71 | 5 | 2 |
| Ngerenya | 75 | 63 | 76 | 67 | 77 | 67 | 78 | 74 | 1 | 5 |
| Roka | 76 | 66 | 75 | 69 | 75 | 67 | 81 | 73 | 3 | 3 |
| Sokoke | 69 | 64 | 79 | 74 | 80 | 71 | 82 | 75 | 6 | 5 |
| Takaungu Mavueni | 70 | 69 | 76 | 72 | 77 | 70 | 79 | 70 | 4 | 1 |
| Tezo | 71 | 63 | 75 | 70 | 76 | 70 | 79 | 73 | 3 | 4 |
| Ziani | 71 | 65 | 77 | 69 | 77 | 67 | 82 | 69 | 5 | 2 |
| **Range** | 13 | 8 | 6 | 7 | 11 | 7 | 5 | 6 |  |  |
| **Standard deviation** | 3 | 3 | 2 | 2 | 3 | 2 | 2 | 2 |  |  |

*Standard deviation of male and female life expectancy across the entire period 2003-18

|  | **2003-06** | | | | **2007-10** | | | | **2011-14** | | | | | **2015-18** | | | |
| --- | --- | --- | --- | --- | --- | --- | --- | --- | --- | --- | --- | --- | --- | --- | --- | --- | --- |
| **Covariate** | **IRR** | **p-value** | **95% CI** | | **IRR** | **p-value** | **95% CI** | | **IRR** | **p-value** | **95% CI** | | | **IRR** | **p-value** | **95% CI** | |
| sex |  |  |  |  |  |  |  |  |  |  |  | |  |  |  |  |  |
| female | 1 | - | - | - | 1 | - | - | - | 1 | - | - | | - | 1 | - | - | - |
| male | 1.17 | 0.000 | 1.11 | 1.23 | 1.25 | 0.000 | 1.19 | 1.32 | 1.39 | 0.000 | 1.31 | | 1.46 | 1.47 | 0.000 | 1.40 | 1.56 |
| Agegroup (yrs) |  |  |  |  |  |  |  |  |  |  |  | |  |  |  |  |  |
| <5 | 1 | - | - | - | 1 | - | - | - | 1 | - | - | | - | 1 | - | - | - |
| 5-14 | 0.11 | 0.000 | 0.10 | 0.12 | 0.11 | 0.000 | 0.10 | 0.13 | 0.14 | 0.000 | 0.12 | | 0.16 | 0.15 | 0.000 | 0.13 | 0.17 |
| 15-54 | 0.44 | 0.000 | 0.41 | 0.47 | 0.51 | 0.000 | 0.48 | 0.55 | 0.52 | 0.000 | 0.48 | | 0.56 | 0.55 | 0.000 | 0.51 | 0.60 |
| 55-74 | 1.95 | 0.000 | 1.81 | 2.09 | 3.23 | 0.000 | 2.99 | 3.49 | 3.64 | 0.000 | 3.37 | | 3.94 | 3.63 | 0.000 | 3.34 | 3.95 |
| ≥75 | 5.90 | 0.000 | 5.40 | 6.45 | 11.03 | 0.000 | 10.11 | 12.02 | 11.82 | 0.000 | 10.83 | | 12.91 | 14.70 | 0.000 | 13.48 | 16.03 |
| **Random effect: location** | **variance** | | **95% CI** | | **variance** | | **95% CI** | | **variance** | | **95% CI** | | | **variance** | | **95% CI** | |
|  | 0.006048 | | 0.0022 | 0.0163 | 0.001976 | | 0.0002 | 0.0142 | 0.001875 | | 0.0002 | 0.0131 | | 0.004495 | | 0.0012 | 0.0161 |

Table S8. Output from the random effect Poisson regression model used to estimate geographical variation in mortality after adjusting for age and sex

Table S9. Infant and under-five mortality ratios from the KHDSS, DHS and KNBS

|  | IMR | | | U5MR | | |
| --- | --- | --- | --- | --- | --- | --- |
| **Kilifi** | 1993-2002 | 1998-2007 | 2004-2013 | 1993-2002 | 1998-2007 | 2004-2013 |
| KHDSS | - | - | 31 | - | - | 46 |
| DHS | 78 | 71 | 44 | 116 | 87 | 57 |
| KNBS* | - | - | 42 | - | - | 57 |

1. KNBS data is for Kilifi county and KHDSS covers a section of Kilifi county

2. DHS data are for entire coast province which includes the whole of Kilifi County and the KHDSS area

3. DHS 1993-02 is obtained from the 2003 report that estimated mortality in the preceding 10 years before the survey in 2003

4. DHS 1998-07 is obtained from the 2008 report that estimated mortality in the preceding 10 years before the survey in 2008

5. DHS 2004-13 is obtained from the 2014 report that estimated mortality in the preceding 10 years before the survey in 2014

6. KHDSS was not fully operational before 2003 so there are no data in these cells

7. *The KNBS estimates are not an average of the entire 2004-13 period but of deaths that occurred in the 12 months preceding the census in 2009

8. KHDSS estimates for the period 2004-13 is the average of the annual mortality rates in the years in that period.

Figure S1. Location of the Kilifi County in Kenya and a map showing the population density of the KHDSS across the 15 administrative locations.

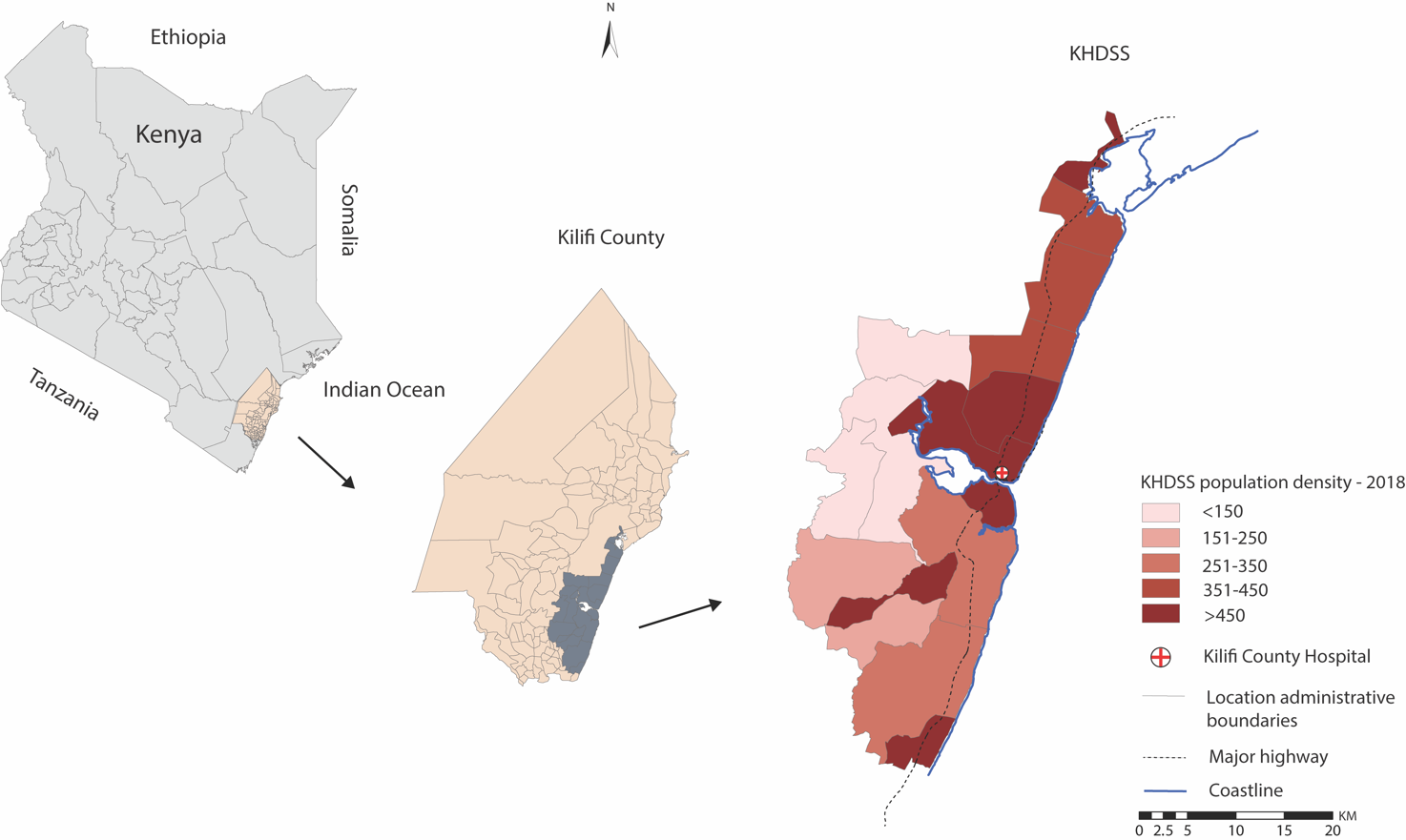

Figures S2a. Age-specific mortality rates (per 1000 PYO) by period

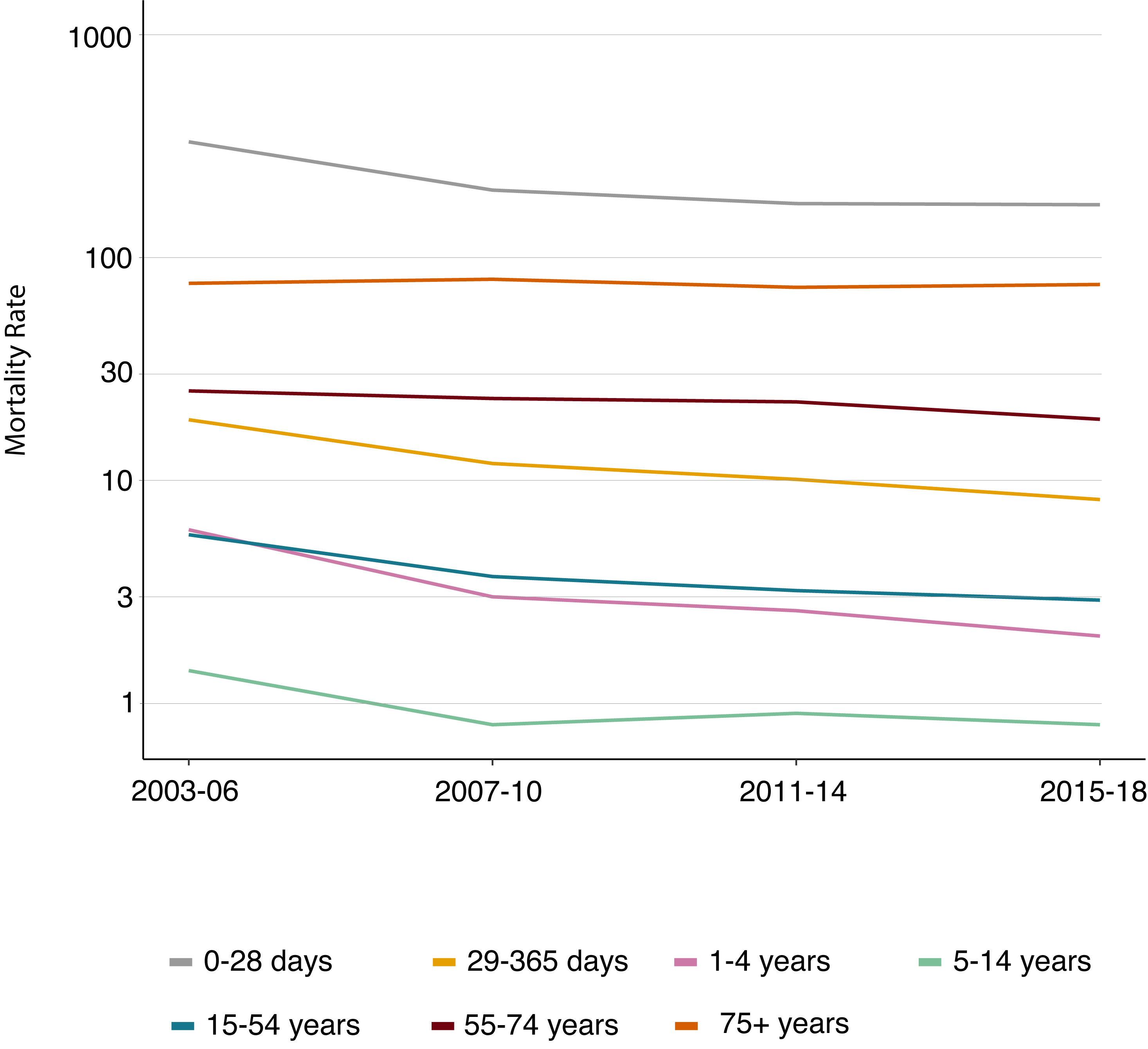

Figures 2Sb. Period mortality rates (per 1000 PYO) by age

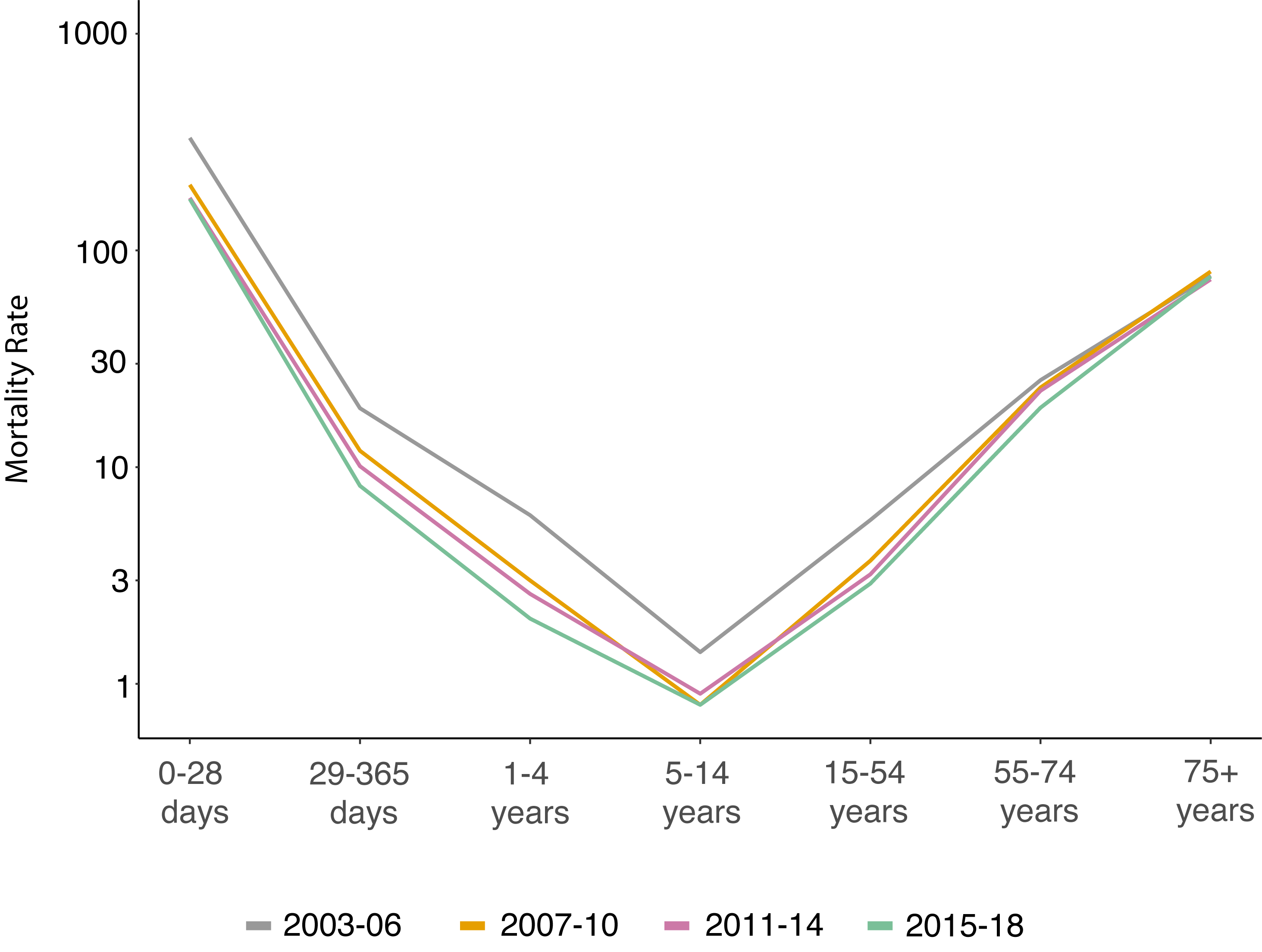

Figures S3a. Monthly mortality rates in neonates over the entire 16-year period. The smoothed trend is estimated from the data series using the LOESS smoothing procedure.

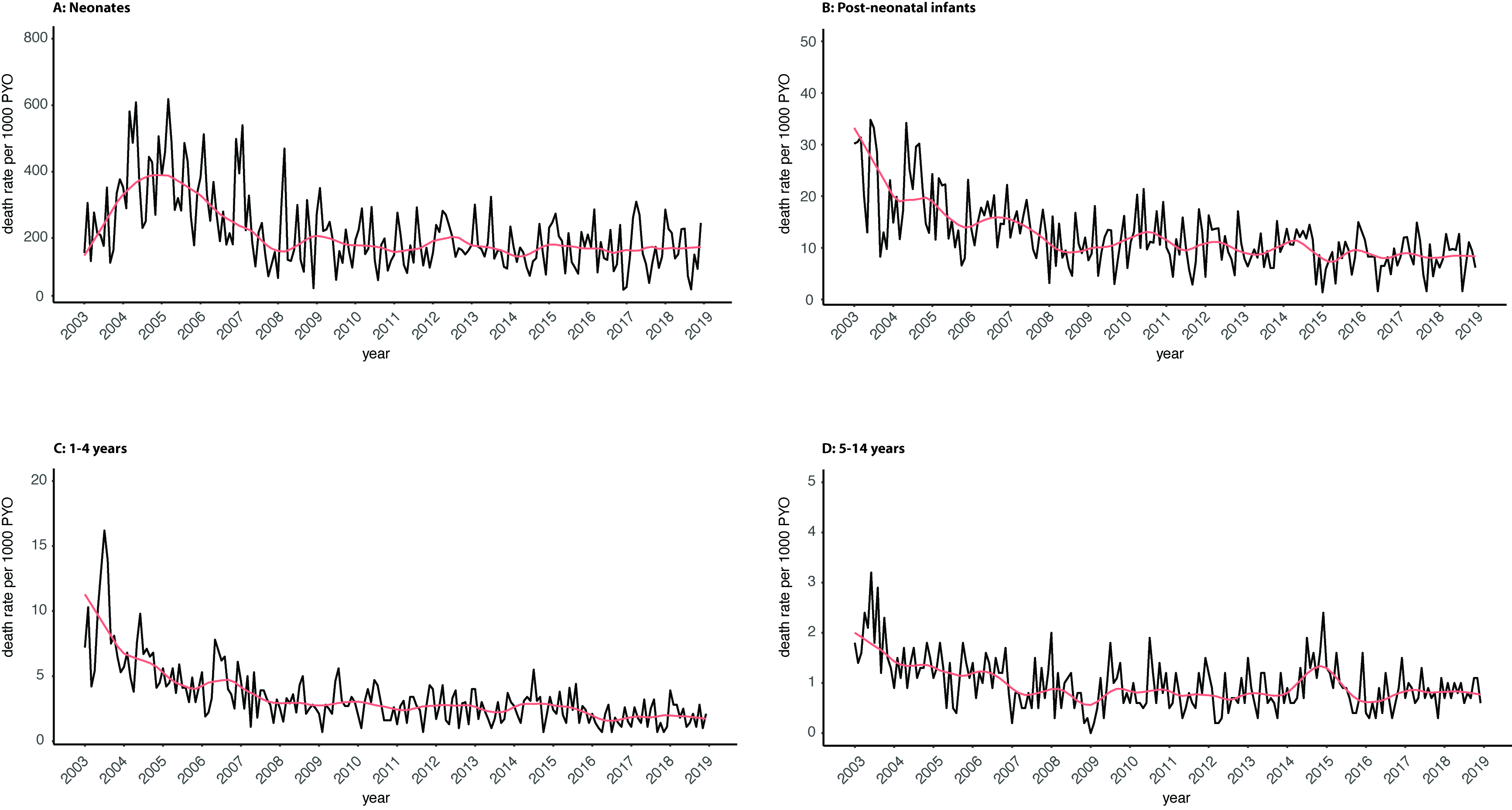

Figures S3b. Monthly mortality rates in neonates over the entire 16-year period. The smoothed trend is estimated from the data series using the LOESS smoothing procedure.

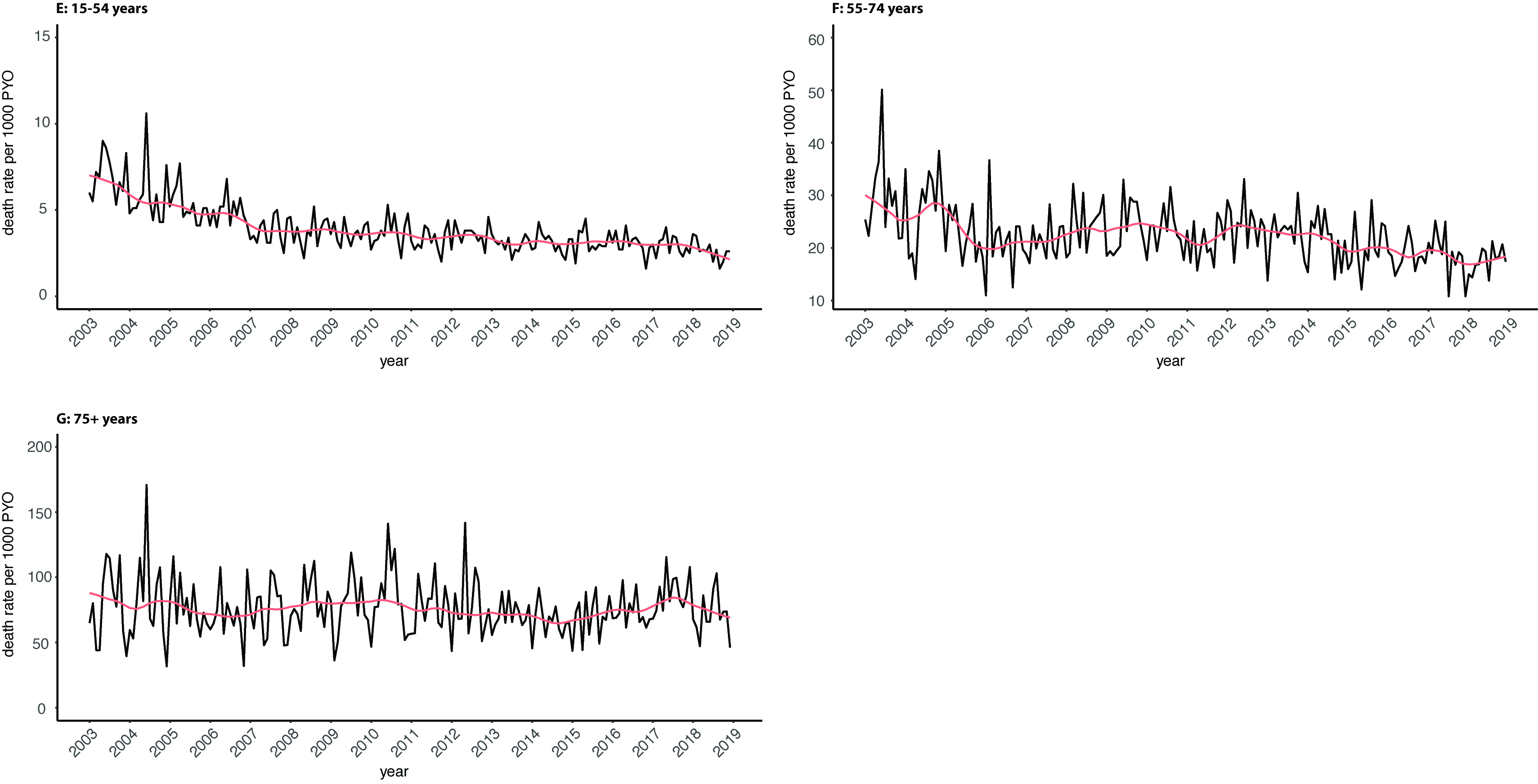

Figures S4a. Mortality rates by location and time-period in different age groups of children

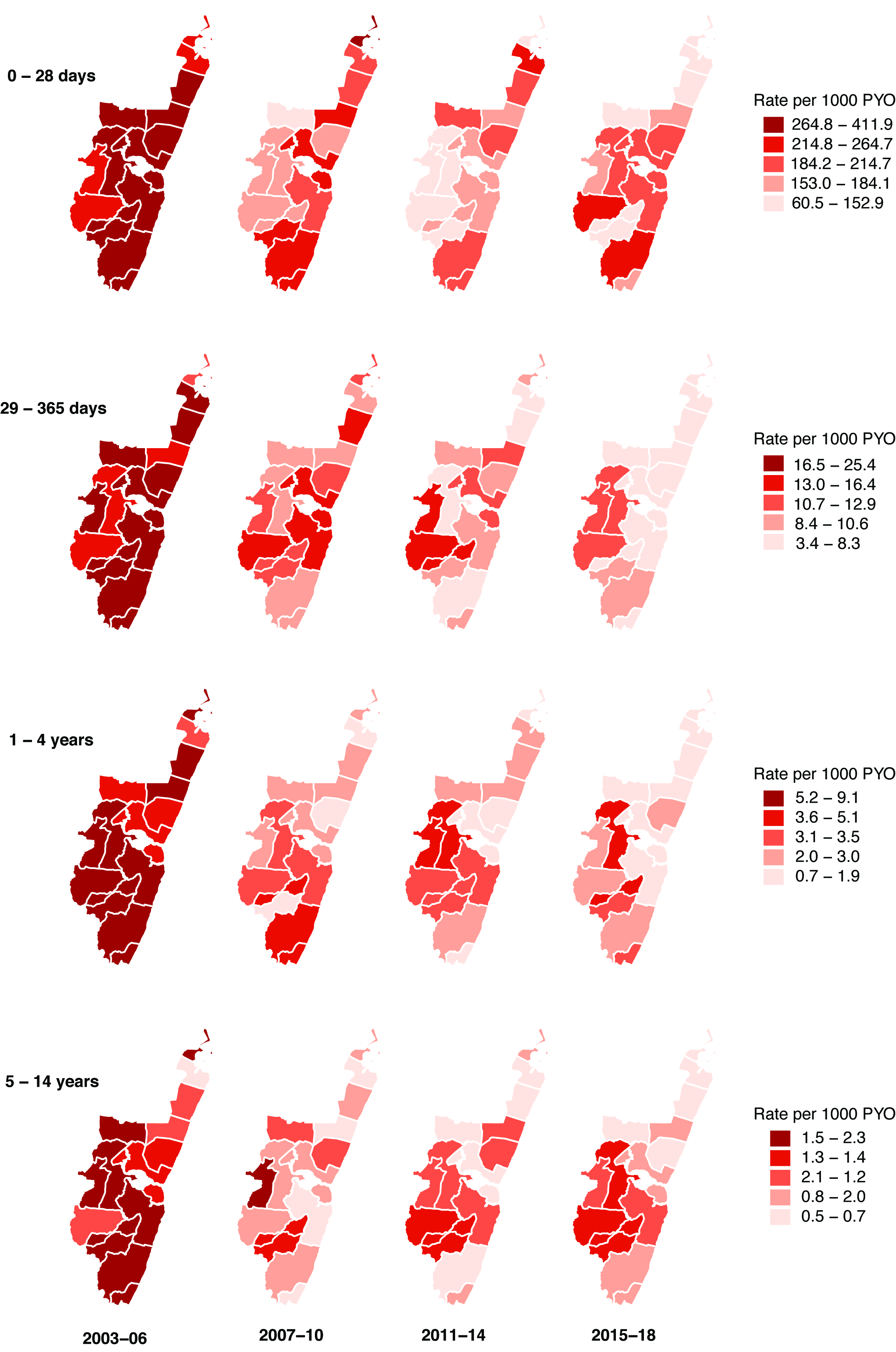

Figures S4b. Mortality rates by location and time-period in different age groups of adults

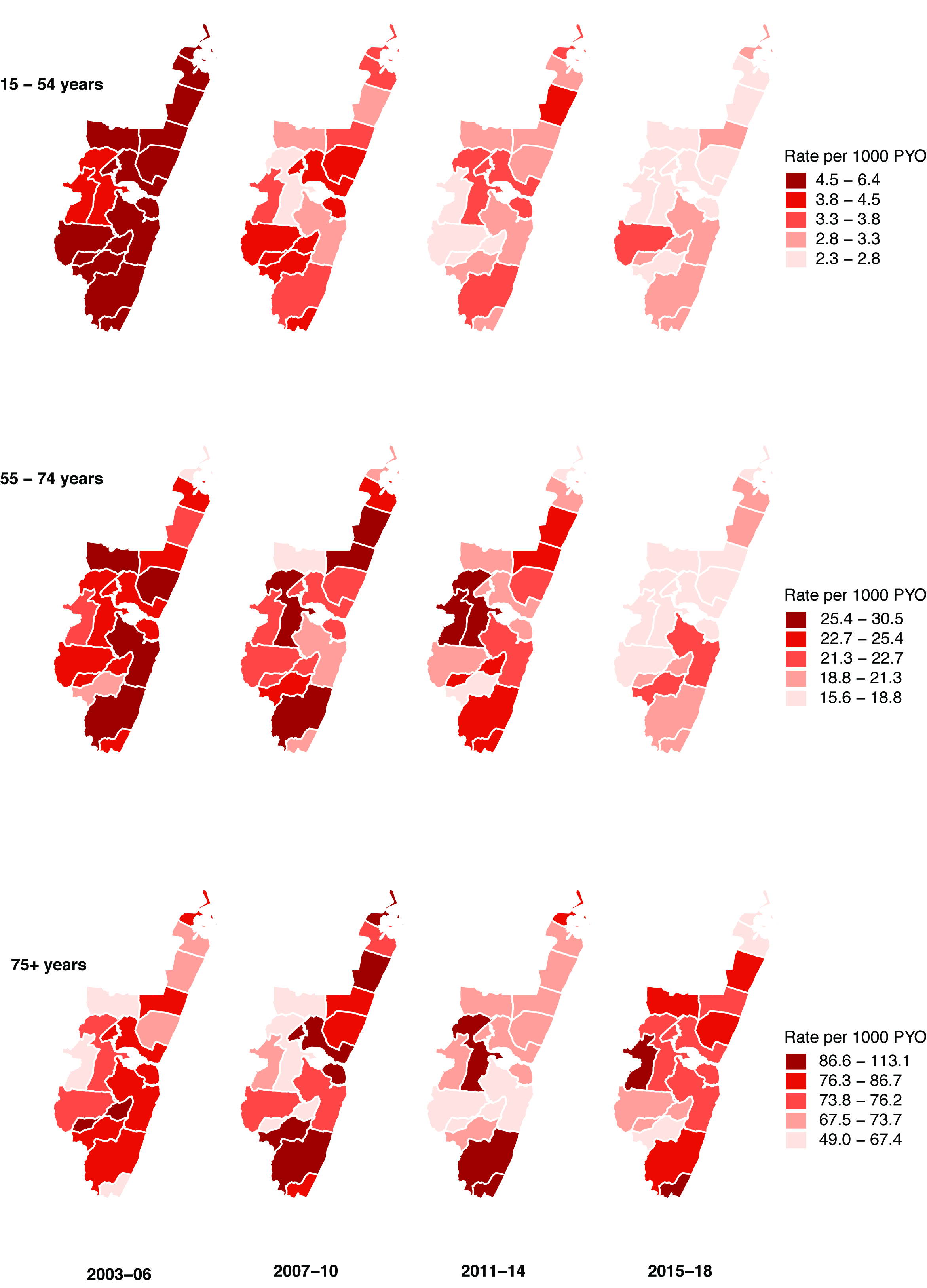

Figures S5. Sex-specific mortality rates by age for each of the 4-year periods

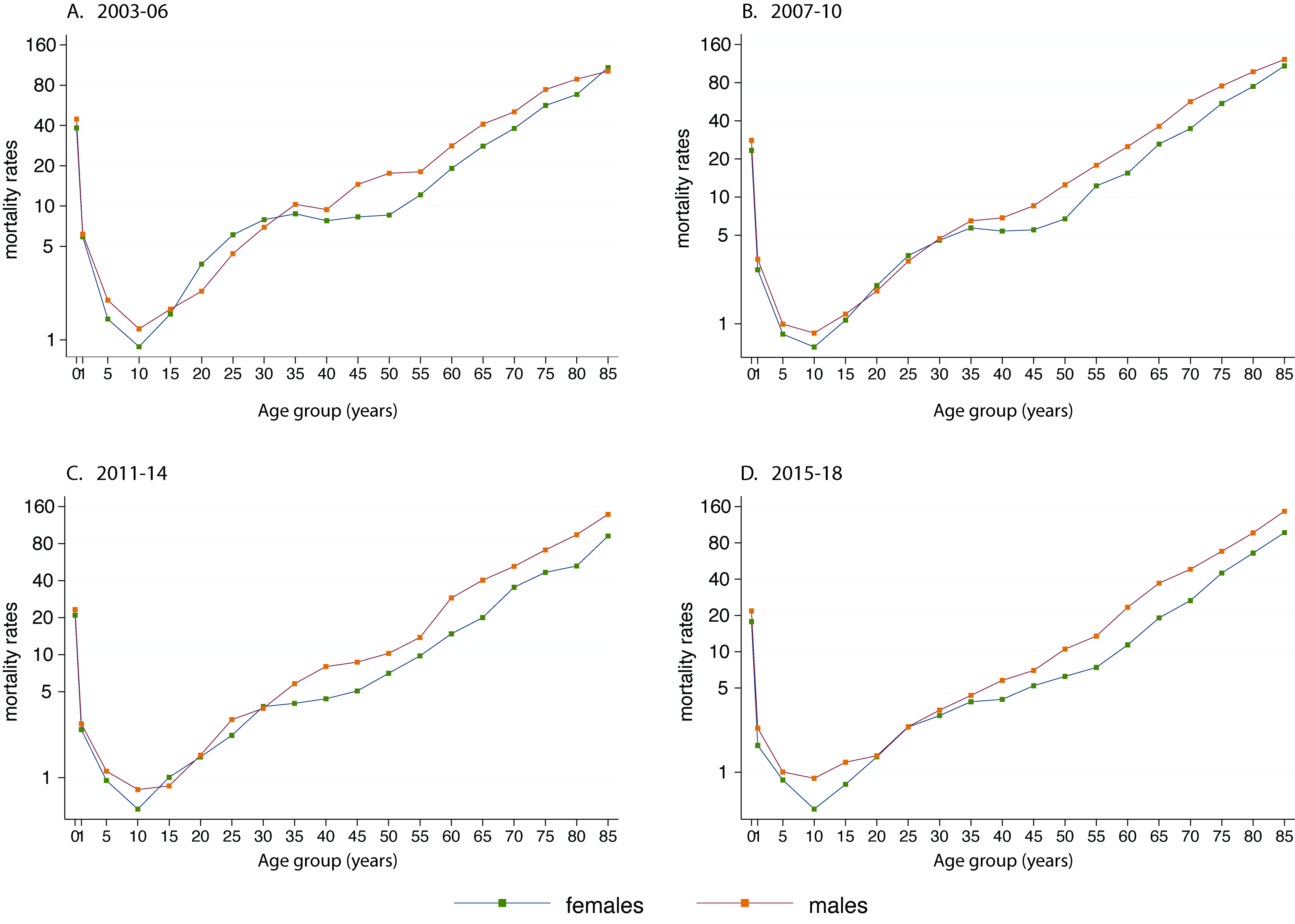
